# Genome-wide association study reveals different T cell distributions in peripheral blood of healthy individuals at high genetic risk of type 1 diabetes and long COVID

**DOI:** 10.1101/2024.02.08.24302520

**Authors:** Laura Deecke, Jan Homann, David Goldeck, Felix Luessi, Marijne Vandebergh, Olena Ohlei, Sarah Toepfer, Frauke Zipp, Ilja Demuth, Sarah L. Morgan, Lars Bertram, Graham Pawelec, Christina M. Lill

**Affiliations:** Institute of Epidemiology and Social Medicine, University of Münster, Münster, Germany; Department of Internal Medicine 2, University of Tübingen, Tübingen, Germany; Fairfax Centre, Kidlington, United Kingdom; Department of Neurology, Focus Program Translational Neuroscience (FTN), Rhine-Main Neuroscience Network (rmn2), University Medical Center of the Johannes Gutenberg University Mainz, Mainz, Germany; VIB Center for Molecular Neurology, University of Antwerp, 2610, Antwerp, Belgium; Department of Biomedical Sciences, University of Antwerp, Antwerp, Belgium; Leuven Brain Institute, Department of Neurosciences, University of Leuven, Leuven, Belgium; Lübeck Interdisciplinary Platform for Genome Analytics (LIGA), University of Lübeck, Lübeck, Germany; Charité – Universitätsmedizin Berlin, corporate member of Freie Universität Berlin and Humboldt-Universität zu Berlin, Department of Endocrinology and Metabolic Diseases (including Division of Lipid Metabolism), Berlin, Germany; Berlin Institute of Health at Charité - Universitätsmedizin Berlin, BCRT - Berlin Institute of Health Center for Regenerative Therapies, Berlin, Germany; Department of Neuroscience, Sheffield Institute for Translational Neurosciences, University of Sheffield, Sheffield, UK; Department of Immunology, University of Tübingen, Tübingen, Germany; Health Sciences North Research Institute of Canada, Sudbury, Ontario, Canada; Ageing and Epidemiology Unit (AGE), School of Public Health, Imperial College London, London, UK

**Author notes:** Corresponding Author: Dr. Christina Lill, MD, MSc, Translational Epidemiology Unit, Institute of Epidemiology and Social Medicine, University of Münster, Domagkstr. 3, 48149 Münster, Germany. equally contributing (Laura Deecke, Jan Homann, David Goldeck).

**Keywords:** GWAS, immune cells, autoimmune disease, polygenic risk score, COVID-19, CD4+, CD8+

## Abstract

The immune system plays a crucial role in many human diseases. In this context, genome-wide association studies (GWAS) offer valuable insights to elucidate the role of immunity in health and disease. The present multi-omics study aimed to identify genetic determinants of immune cell type distributions in the blood of healthy individuals and to assess whether the distributions of these cells may play a role for autoimmune and COVID-19 disease risk.

To this end, the frequencies of different immune cells in 483 healthy individuals from the Berlin Aging Study II were quantified using flow cytometry, and GWAS was performed for 92 immune cell phenotypes. Additionally, we performed linear regression analyses of immune cell distributions using polygenic risk scores (PRS) based on prior GWAS for five autoimmune diseases as well as for COVID-19 infection and post-COVID syndrome (“long COVID”).

We validated seven previously described immune loci and identified 13 novel loci showing genome-wide significant (α=5.00E-8) association with different immune cell phenotypes. The most significant novel signal was conferred by the *SLC52A3* locus, encoding for a riboflavin transporter protein, which was associated with naïve CD57+ CD8+ T cells (p=4.13E-17) and colocalized with *SLC52A3* expression. Several novel loci contained immunologically plausible candidate genes, e.g., variants near *TBATA* and *B3GAT1* representing genes associated with T cell phenotypes. The PRS of type 1 diabetes were significantly associated with CD8+ T cells at different differentiation states (p≤7.02E-4), and PRS of long COVID were associated with early-differentiated CD4+ T cells (p≤1.54E-4).

In conclusion, our extensive immune cell GWAS analyses highlight several novel genetic loci of likely relevance for immune system function. Furthermore, our PRS analyses point to a shared genetic basis between immune cell distributions in healthy adults and T1D (CD8+ T cells) as well as long COVID (CD4+ T cells).

## Introduction

It is becoming increasingly evident that the immune system plays a crucial role not only in infectious and inflammatory diseases but also in a wide range of other human disorders, including cardiovascular diseases, cancer, neurological and even psychiatric diseases^1,2^. In this context, genome-wide association studies (GWAS) offer the possibility of elucidating physiological and pathophysiological mechanisms of immune system function and of investigating the genetic overlap of immune system-related phenotypes and human diseases^3,4^. At the same time, in-depth immune phenotyping using multicolor flow cytometry now allows comprehensive investigation of the composition of immune cell types in the blood. This blood-based immune cell signature reflects the impact of genetic, environmental, and lifestyle factors, and may reflect predisposition to disease^3,5,6^. Accordingly, in recent years, several GWAS have already been performed on immune cell type compositions in blood samples of healthy adults and have identified several dozen potential immune cell loci^3,7–10^ (**Figure 1**, **Supplementary Table 1**). Fourteen such immune cell loci were described in at least two independent studies, including the Fc gamma receptor (*FCGR*) gene locus on chromosome 1q23.3^3,7,8,10^, the *MIR181A1HG*/*PTPRC* locus on chromosome 1q31.3-q32.1^7,9^ and the *ANO2* locus on chromosome 12p13.31^7,10^ (**Figure 1**). In those studies, heritability of immune cell type compositions calculated from genome-wide genotyping data was estimated to be nearly 40% (median; range ∼30-50%). Notably, the most heritable cell types tended to be cells of the adaptive immune system, with complex functions^7^. These heritability estimates suggest that other non-heritable factors also impact immune cell type composition in blood. This has been described for several intrinsic and environmental factors, including age^3,10–12^, sex^3,10^, cytomegalovirus (CMV) infection^3,12^, and smoking^3^.

**Figure 1.**
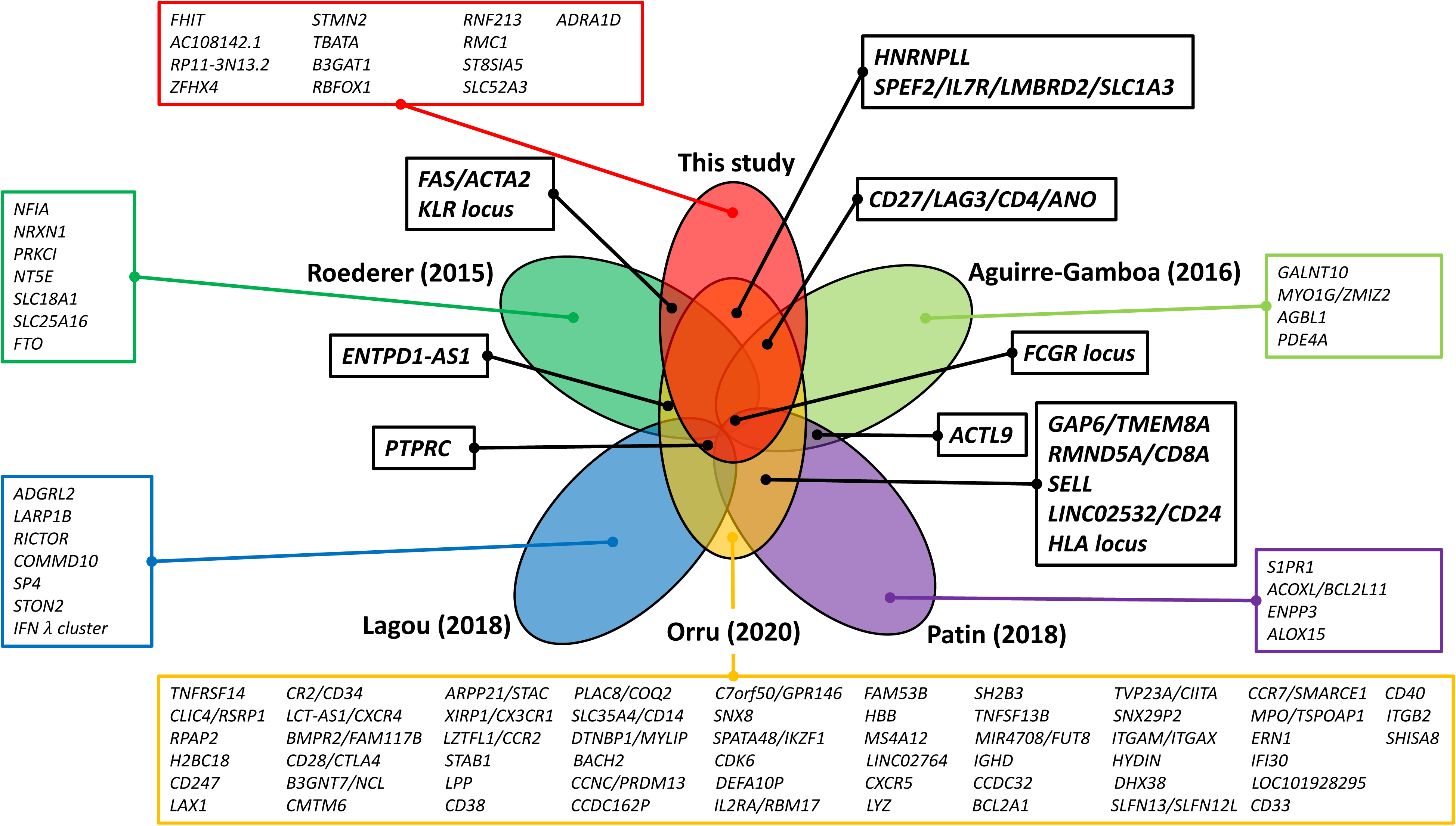
Overlap between immune cell loci identified by this study and by most recent genome-wide association studies in the field. This Venn diagram displays all genetic loci and SNPs that were reported to show genome-wide significant association with immune cell types in this study and in relevant publications^3,7–10^. Association signals were generally assigned to the gene names used in the original publications; association signals within 1Mb were merged. For association signals assigned to more than one gene name, one gene name was selected for better legibility.

Here, we performed GWAS analyses on 92 different immune cell phenotypes in 483 immunologically healthy individuals from the Berlin Aging Study II (BASE-II) and integrated these data with large-scale data from other omics domains (e.g., transcriptomics). In addition, we investigated previously described associations of age, sex, CMV infection, and smoking status with immune cell compositions. Finally, by analyzing the impact of polygenic risk scores (PRS) on the immune cell data, we assessed whether autoimmune disease, COVID-19, and post-COVD syndrome (“long COVID”) predisposition are associated with differential immune cell distributions in healthy individuals before disease manifestation.

## Methods

A detailed description of the methods can be found in the **Supplementary Material**.

### Study participants

In this study, we investigated 483 immunologically healthy participants of European descent from the BASE-II project, a multi-institutional and multi-disciplinary longitudinal study aimed at investigating factors modulating the aging process in inhabitants of the greater metropolitan area of Berlin, Germany^13,14^. All participants were part of the baseline recruitment of BASE-II and comprised a group of 344 older (aged 60-82 years) and 139 younger adults (aged 23-35 years, **Supplementary Table 2)** for whom quality-controlled genetic and immune cell data were available. As the aim of the study was to identify determinants of immune cell composition in healthy individuals, we included only those BASE-II participants without prevalent immune system-related diseases or treatments at baseline (see **Supplementary Methods**).

### Generation of immune cell data by flow cytometry

Isolation of peripheral blood mononuclear cells (PBMCs) from whole blood samples and subsequent flow cytometry was performed as previously described^15,16^: Briefly, PBMCs were isolated, cryopreserved and biobanked in liquid nitrogen. For flow cytometry, PBMCs were thawed and two different antibody panels were applied, which comprised various differentiated T lymphocytes (panel 1), monocytes, natural killer (NK) cells, natural killer T (NKT) cells, myeloid-derived suppressor cells (MDSC), and B cells (panel 2; **Supplementary Figure 1**, **Supplementary Table 3**). Specifically, after thawing, PBMCs were incubated with GAMUNEX (human IgG; Bayer, Leverkusen, Germany) and ethidium monoazide (EMA) bromide (MoBiTec GmbH, Göttingen, Germany). For panel 1, cells were then incubated with mouse anti human CCR7 (CD197; R&D Systems, Minneapolis, Minnesota, USA), anti-mouse IgG Pacific Orange (Invitrogen, Waltham, Massachusetts, USA) and mouse serum. Next, for both panels, cells were stained with monoclonal antibodies against the markers listed in **Supplementary Table 4.** The cells were acquired with a 3 laser BD LSRII (BD biosciences, Heidelberg, Germany) flow cytometer and DIVA6 software. Samples were analyzed with FlowJo version 7.5 (TreeStar, Portland, USA). Immune cells were analyzed as proportions of parental immune cell populations in blood, and where applicable, following visual inspection, immune cell proportions were transformed using log, root, log(100-x) and square transformations, respectively (**Supplementary Figure 2, Supplementary Table 3**).

Some of the immune cell proportions analyzed in this study correlated with each other. Thus, we performed hierarchical clustering (calculated with R package NbClust^17^ [ward method, Euclidean distance]). Where appropriate, we present the results by clusters. The optimal number of clusters that best represents the data structure was determined by the majority rule on a combination of cluster analysis methods implemented in NbClust.

### Generation of genome-wide SNP data

Genome-wide microarray-based genotyping on blood DNA was performed using the Genome-Wide Human SNP Array 6.0 (Affymetrix Inc., Santa Clara, California, USA). Data processing and analysis were performed with PLINK v1.9 (www.cog-genomics.org/plink/1.9/) or v2.0 (www.cog-genomics.org/plink/2.0/)^18^, unless stated otherwise. Prior to imputation, the genotype data underwent standard quality control (QC) as previously described^16,19^ (**Supplementary Methods**). Next, we performed genotype imputation on 723,727 genotyped and QC’ed SNPs using the Haplotype Reference Consortium reference panel and Minimac3 as described previously^19^ resulting in a total of 38,407,851 SNPs. For post-imputation QC, the same QC criteria were applied as for the pre-imputation QC. In addition, we excluded SNPs with an imputation quality score R^2^<0.7. Following QC and merging of the genetic and phenotypic data, the effective sample size included in all subsequent statistical analyses was 483 participants (420 individuals for panel 1 and 352 individuals for panel 2; **Supplementary Table 3** for exact sample sizes per GWAS analysis) and 6,932,885 SNPs.

### Statistical analyses on immune cell distributions

GWAS were performed on 51 immune cell phenotypes from panel 1 and 41 phenotypes from panel 2 using linear regression analyses adjusting for sex and principal components (PC) 1-4 to account for ancestry. GWAS for each immune cell subpopulation were run separately for both age groups and subsequently combined by fixed-effect meta-analyses. Heterogeneity of effects was estimated using the /^2^ statistic. Possible inflation of the GWAS test statistics was assessed by visual inspection of the corresponding quantile-quantile (QQ) plots and by calculation of the respective inflation factor λ^20^. We visualized the main results in a Manhattan plot generated by the R package CMplot^21^, which allowed us to display all GWAS results in one plot by color-coding the different cell type clusters (“Candy plot”). Genome-wide significance was defined at an α=5.00E-8. To identify additional independent signals for each locus with a genome-wide significant association signal, we performed linear regression analyses conditioning on the index SNP in a ±1Mb window. The conditional results were false discovery rate (FDR)- controlled at 1%. In addition, we examined the previously reported (see above) association of age (older vs younger age group), CMV status (positive vs negative), sex (women vs men), and smoking (current vs non-current [former/never smokers]), and the 92 immune cell phenotypes using multivariate linear regression (lm function in R). The results were FDR-controlled at 1%.

### Functional annotations of GWAS results

Index SNPs and, where applicable, their proxies (r^2^>0.6) were annotated according to genomic location, nearest gene, functional consequence, Combined Annotation–Dependent Depletion (CADD) score^22^, molecular *cis* quantitative trait loci (QTL) effects, and colocalization analyses^23^. *Cis* QTL effects were assessed using the QTLbase^24^ (http://www.mulinlab.org/qtlbase/index.html, downloaded on March 16th, 2022). Specifically, we considered *cis* QTL results for four molecular markers (i.e., expression QTL [eQTL], protein QTL [pQTL], methylation QTL [mQTL] and histone modification QTL [hQTL]) in whole blood and in a selection of 12 blood cell types depending on their relevance to the respective GWAS finding (i.e., B cells, monocytes, CD14+ monocytes, NK cells, CD16+ neutrophils, CD4+ T cells, naïve CD4+ T cells, CD8+ T cells, activated CD8+ T cells, naïve CD8+ T cells, and lymphocytes). We considered all molecular QTLs with p<1.00E-05. We assessed a shared genetic etiology for both the immune cell GWAS signal and gene expression signals in the same locus by performing colocalization analyses^23^ of each GWAS region (index SNP ±1Mb) using the largest blood eQTL dataset published to date^25^ (n=31,684; data downloaded from QTLbase on May 04^th^, 2022) based on the R package *coloc* v5 (https://cran.r-project.org/web/packages/coloc/index.html)^26^ (**Table 1, Supplementary Table 5**). Colocalization of association signals was defined as the posterior probability of a shared causal variant >80% (‘PP.H4’>0.8). Furthermore, we compared our genome-wide significant findings with those from previous immune cell GWAS^3,7–10^ (and vice versa) by using the data provided in the original publication^3,7,8,10^ or by using GWAS summary statistics made available to us by the investigators^9^ (**Table 1, Supplementary Table 1**), and to immunologically relevant phenotypes listed in the GWAS Catalog^27^.

**Table 1.**
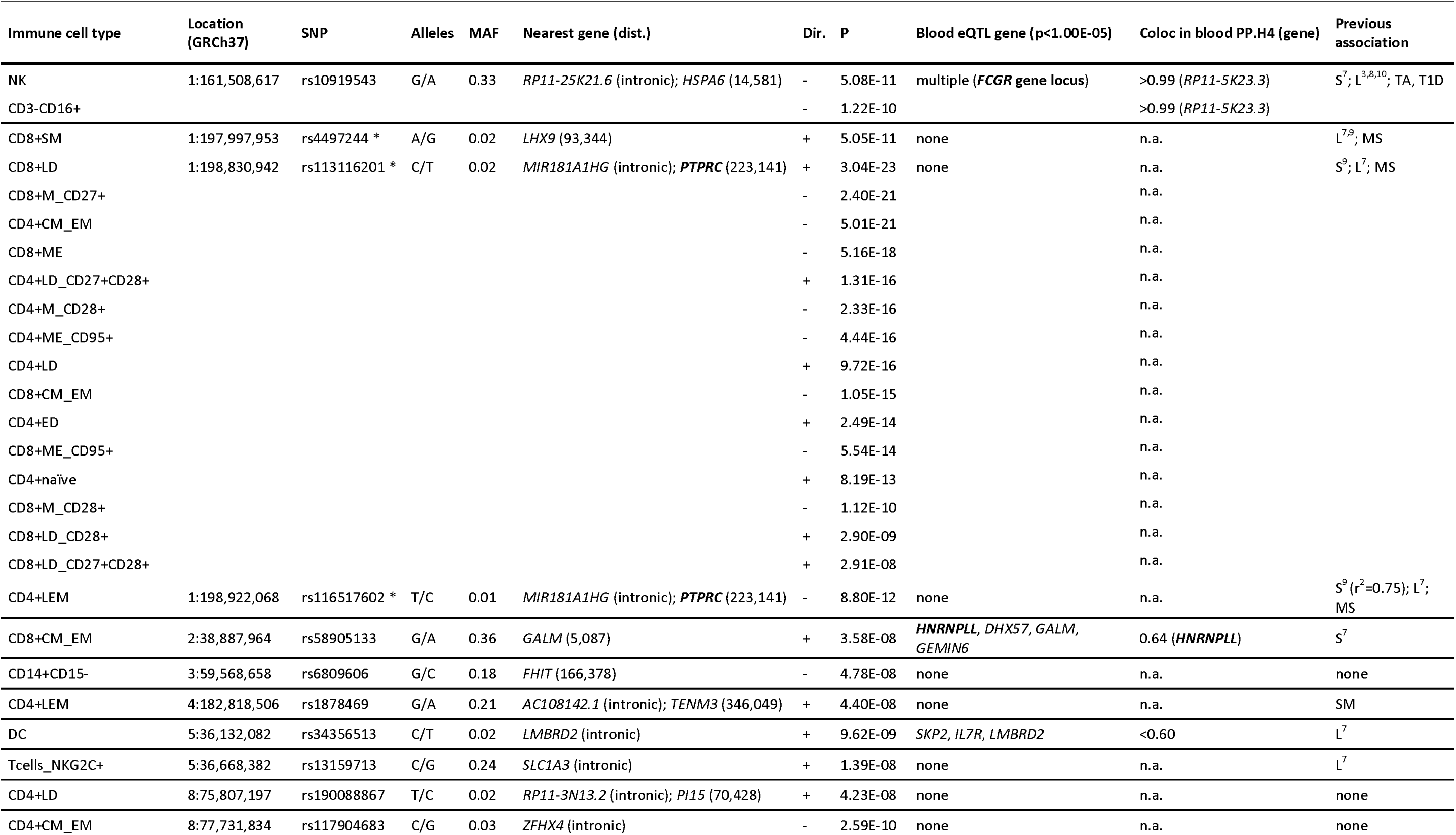
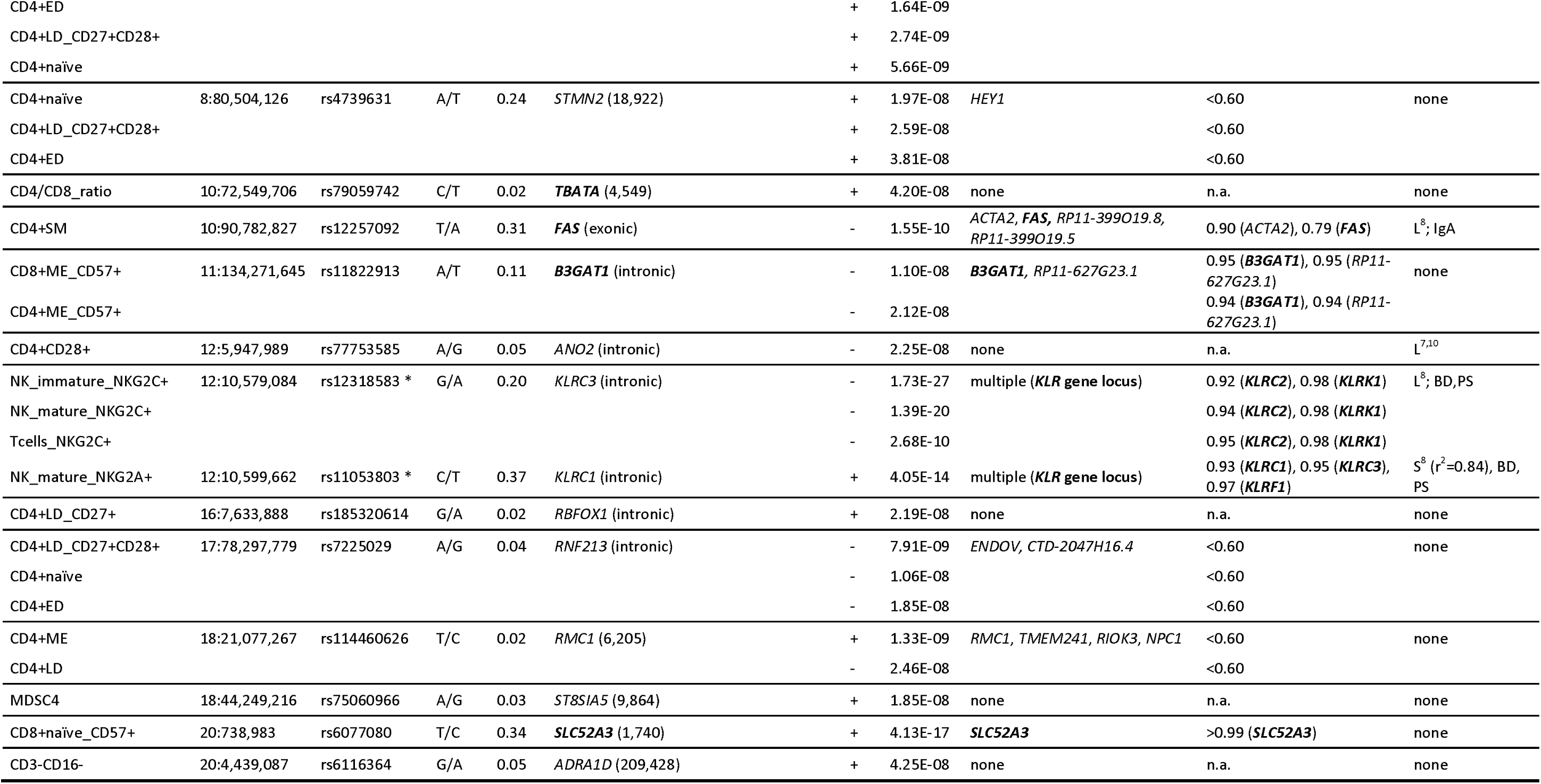
Genome-wide significant association results for immune cell types assessed in 483 BASE-II participants. This table lists all independent index SNPs (r <0.1, exceptions are marked with “*”) showing genome-wide significant (α=5.00E-8) association with immune cell types quantified in 483 BASE-II participants. The full description of the immune cell phenotypes displayed (“Immune cell type”) can be found in Supplementary Table 3. Bold gene names indicate compelling candidate genes upon review of all available data. Please note that rs4497244, rs113116201 and rs116517602 as well as rs12318583 and rs11053803 (labeled with “*”) are not independent (r >0.3) from the other index SNPs in the same locus but represent the index SNPs for distinct immune cell phenotypes. dist.=distance (in bp) to the nearest gene using ANNOVAR ; alleles=minor/major allele; MAF=minor allele frequency; dir=direction of effect estimate for the minor allele; P=p-value; “Blood eQTL gene” lists all blood-based expression quantitative trait loci (eQTL) with p<1.00E-05; note that for the immunological complex FCGR and KLR loci the entry ‘multiple’ is displayed due to the large range of possibly functional genes (Supplementary Table 8); Coloc=colocalization results of the blood eQTL and the BASE-II immune cell datasets with PP.H4>0.6; previous association=reported in previous GWAS on immune cells or and/or immunologically relevant diseases; S=same SNP as in this study or SNP in high LD (r >0.6) reported for immune cells, L=same locus (+/-1Mb window) reported for immune cells; a disease name is listed if the immune cell index SNPs or SNPs in LD (r2>0.6) are listed in the GWAS catalog for the specific disease. TA=takayasu arteritis, T1D=type 1 diabetes (age at diagnosis), MS=multiple sclerosis, IgA=Selective IgA deficiency, BD=Behcet’s disease, PS=psoriasis, SM=severe malaria.

### Polygenic risk score analyses

Next, we assessed whether autoimmune diseases or COVID-19 phenotypes already manifest themselves by differential immune cell type compositions in healthy adults. Using PRSice-2, we calculated the best-fitting PRS on five autoimmune diseases and on COVID-19: These analyses based on large, publicly available GWAS summary statistics on multiple sclerosis^29^, type 1 diabetes (T1D)^30^, rheumatoid arthritis^31^, Crohn’s disease^32^, ulcerate colitis^32^, COVID-19 (three partly overlapping case-control GWAS datasets)^33^, long COVID^34^ (four largely similar datasets using strict or broader case and control definitions), and – as a negative control – human height^35^. We performed linear regression analyses of the 92 immune cell types in the BASE-II dataset on the respective PRS adjusting for sex and the first four PCs. Empirical p values were derived using 10,000 permutations (--perm 10000). We chose a conservative approach and defined a result as being statistically significant if it showed empirical significance (α=0.01) following permutation *and* if it passed FDR control at 1%. Further, for statistically significant PRS associations in our primary analyses we re-calculated the PRS using only genome-wide significant SNPs from the respective GWAS summary statistics (only applicable to the T1D GWAS summary statistics^30^ due to the absence of multiple genome-wide significant hits in the long COVID summary statistics^34^).

## Results

### Dataset

The effective dataset analyzed in this study comprised 483 immunologically healthy BASE-II participants on whom we performed genome-wide association analyses for 92 immune cell phenotypes across 6,932,885 SNPs (**Supplementary Tables 2-3**). Hierarchical clustering of the 92 immune cell phenotypes in BASE-II yielded 20 clusters (**Supplementary Figure 3**). The correlation matrix revealed many known interdependencies, e.g., a positive correlation between effector memory T cells re-expressing *CD45RA* (*TEMRA*) and CD57+ T cells, a negative correlation between TEMRA cells and early-differentiated T cells^36^ as well as correlations between multiple differentiation states of CD4+ and CD8+ T cells^37^. However, to the best of our knowledge, a few interdependencies were identified that had not been described previously, such as the correlation between NKT cells, MDSCs, and monocytes, as well as the negative correlation between T cell subsets and mature NK cells (**Supplementary Figure 3**).

### Genome-wide association analyses

Genome-wide association analyses on the 92 immune cell types yielded 24 index SNPs in 20 genetic loci that were genome-wide significantly (α=5.00E-8) associated with at least one immune cell type (**Table 1**, **Figure 2).** In three of these 20 loci, the GWAS yielded different index SNPs for different immune cell types (**Table 1**). However, in two of the three loci, these different index SNPs were in at least moderate LD (i.e., rs4497244, rs113116201 and rs116517602 on chromosome 1q with r^2^=0.33-0.75; rs12318583 and rs11053803 on chromosome 12p with r^2^=0.37) possibly pinpointing the same underlying functional genetic variant. Only the two index SNPs on chromosome 5p13.2 (rs34356513, rs13159713) were independent of each other in our dataset (r^2^=0.0009) indicating that they reflect independent functional genetic variants (**Table 1**). A total of 35 of the 92 GWAS (with immune cell types originating from 16 of the 20 cell clusters) yielded at least one genome-wide significantly associated SNP. Furthermore, 14 additional loci (non-overlapping with the 20 genome-wide significant loci) showed borderline genome-wide significant association (p<1.00E-7 but p≥5.00E-8; **Supplementary Tables 6-7**). QQ plots and inflation factor λ did not show any noteworthy inflation of the test statistics in any of the GWAS (λ=0.99-1.03; **Supplementary Figure 4**). The MAF of the index SNPs ranged from 0.01 to 0.37 with approximately half of the SNPs being common with MAF ≥0.05, and half being infrequent with MAF 0.01-0.05. Four loci (*MIR181A1HG/PTPRC*, *ZFHX4*, *KLR* gene locus, *RMC1*) were each associated with immune cell distributions from different clusters (**Table 1**). In addition, the majority of genome-wide significantly associated loci showed sub-genome-wide statistical evidence for association with multiple other immune cell types, originating predominantly from the same cell clusters (**Supplementary Figure 5**). However, a few index SNPs showed strong associations with several immune cell types that did not correlate with each other (r<0.1) suggesting pleiotropic effects: Specifically, the locus on chromosome 1q (rs113116201, *MIR181A1HG/PTPRC*) was associated with CD8+SM cells (cluster 6, p=1.24E-9) and with several independent (r<0.1) CD4+ and CD8+ T cell types, e.g., CD8+CM EM (cluster 9, p=1.05E-15, inverse direction of effect compared to CD8+SM cells), and CD4+ naïve T cells (cluster 19, p=8.19E-13, same direction of effect compared to CD8+SM cells). Furthermore, rs11053803 (*KLRC1* locus) was associated with NK mature NKG2A+ cells (cluster 3, p=4.05E-14) but also showed an independent (r<0.1) inverse suggestive association with NK mature NKG2C+ cells (cluster 13; p=4.87E-6; **Supplementary Figure 5**). Association analyses conditioning on the most significant SNP in each of the genome-wide significantly associated loci yielded one potential second association signal in the locus on chromosome 12p (*KLRC1*). Specifically, the minor allele (T) of rs7309785 (r^2^=0.05 with the index SNP rs12318583) showed an independent significant (FDR=0.01) increase in NK immature NKG2C+ cells (p_uncond_=1.1E-10, p_cond_=2.2E-7). Comparisons of our data with recent immune cell GWAS^3,7–10^ revealed that seven of our 20 genome-wide significantly associated loci had been described at genome-wide significance previously, while 13 loci (*FHIT*, *AC108142.1*, *RP11-3N13.2*, *ZFHX4*, *STMN2*, *TBATA*, *B3GAT1*, *RBFOX1*, *RNF213*, *RMC1*, *ST8SIA5, SLC52A3, ADRA1D*) were novel (**Figure 1**, **Table 1, Supplementary Table 1**). The most significant novel signal was the *SLC52A3* locus that was associated with naïve CD57+ CD8+ T cells (**Table 1**). Furthermore, comparisons with the GWAS Catalog (https://www.ebi.ac.uk/gwas/) implicated several of our index SNPs in a disease with immune system involvement (**Table 1**).

**Figure 2.**
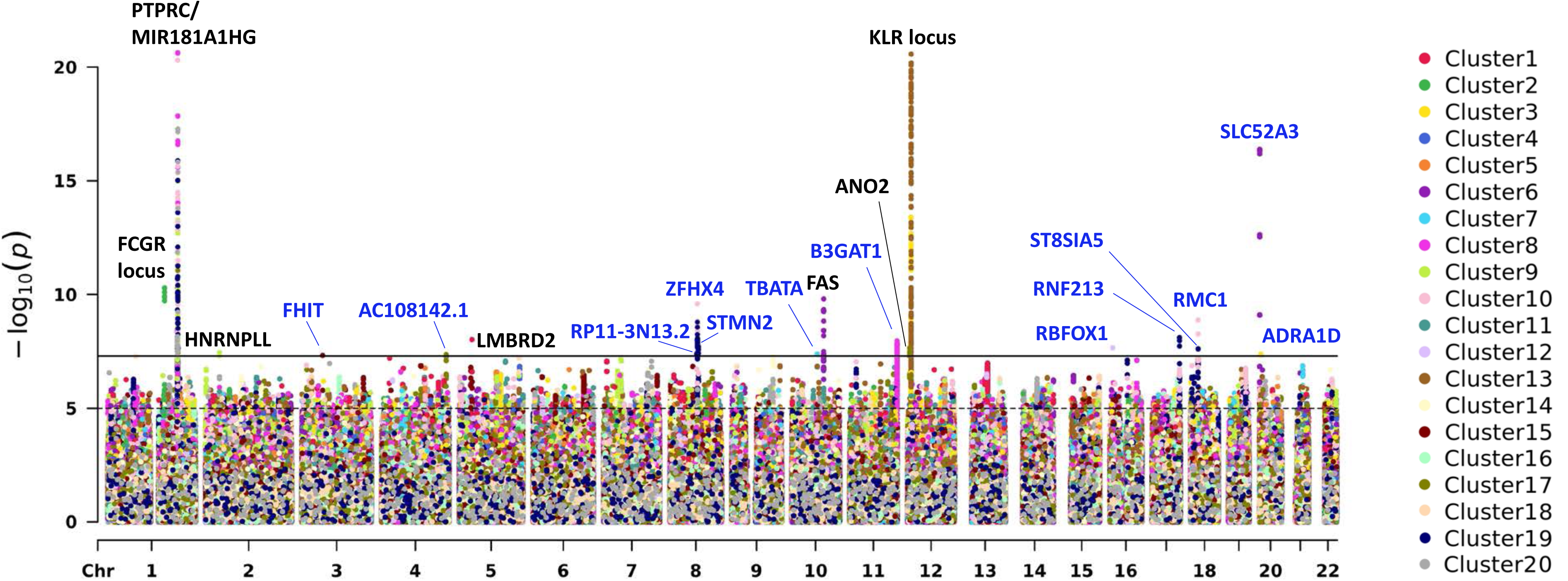
Candy plot of genome-wide association results for 92 immune cell types in the Berlin Aging Study II. This Manhattan-style “Candy plot” shows the genome-wide association results of all 92 immune cell types quantified in 483 BASE-II participants. Clustered immune cell types (**Supplementary Figure 3**) are coded with the same color as indicated in the Figure. For an overview of the immune cell types in each cluster, see **Supplementary Table 3** and **Supplementary Figure 1**. Gene names colored in blue represent novel loci, gene names in black represent previously described loci.

### Molecular quantitative trait locus effects and colocalization analysis

None of the 24 index SNPs was a non-synonymous coding variant. Notably, rs12257092 (associated with CD4+ memory stem T cells in total CD4+ T cells) was located in the 3’ UTR of *FAS* (encoding Fas cell surface death receptor)^38^ (**Figure 3**), which is a member of the TNF-receptor superfamily and functionally active specifically in T cells (NCBI gene ID 355, https://www.ncbi.nlm.nih.gov/gene/). Intriguingly, this SNP showed *FAS* eQTL effects in CD4+ naïve T cells (**Supplementary Table 8**). The other index SNPs were either intergenic (n=10) or intronic (n=13) without any clear evidence for a location near a splice site (also see locus zoom plots in **Supplementary Figure 6**). Twenty-one index SNPs (i.e., all except rs185320614, rs13159713, rs4739631) were in strong LD (r^2^>0.6) with possibly regulatory variants (**Supplementary Table 9**). Next, we investigated the effects of the 24 index SNPs on gene expression (*cis* eQTLs) and other molecular phenotypes (*cis* molQTLs) using whole blood (n=31,684 samples) or relevant blood-based immune cell datasets (n=106 to 696 samples) available in QTLbase. A total of 11 index SNPs (46%) represented one or more significant (p<1.00E-5) molecular QTL in blood including eQTL (n=11 SNPs), pQTL (n=1), and mQTL (n=6), (**Table 1**, **Supplementary Table 8**). The eQTL results on the smaller immune-cell expression datasets did not reveal any additional compelling candidate genes beyond those nominated based on the blood eQTL data (**Supplementary Table 8**). Of all 11 SNPs showing *cis* eQTL effects in blood, the vast majority (n=9) were associated with expressional changes of more than one gene. For instance, rs10919543 on chromosome 1q23.3 was associated with expression levels of 14 genes, and rs58905133 on chromosome 2p22.1 with expression levels of four genes. For eight of the 11 SNPs (73%), the gene located nearest to the index SNP was also the gene highlighted by the eQTL results. The blood-based eQTL results revealed several immunological candidate genes: Apart from several genes from the *FCGR* and *KLR* gene clusters, these comprised *HNRNPLL*, *FAS*, *B3GAT1*, and *SLC52A3* (**Table 1**). Notably, for the latter four genes, eQTL effects of the index SNPs could also be observed in the (compared to the blood eQTL dataset) much smaller but more specific immune cell datasets that are comparable to the cell types for which the original genetic associations with the index SNP were observed (**Supplementary Table 8**).

**Figure 3.**
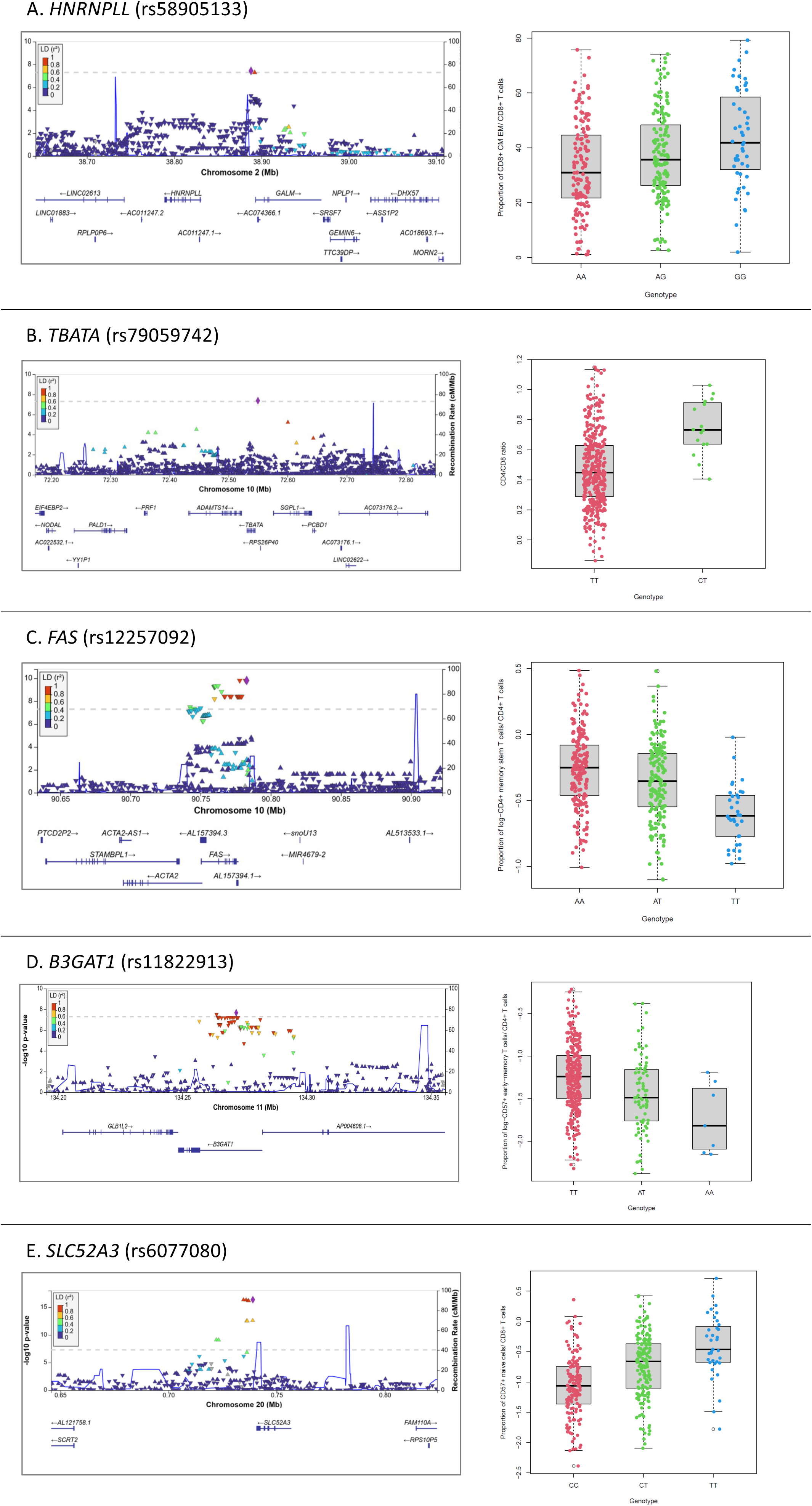
Association results of HNRNPLL rs58905133, TBATA rs79059742, FAS rs12257092, B3GAT1 rs11822913 and SLC52A3 rs6077080. This figure shows locus zoom plots and box and whisker plots for *HNRNPLL* (A), *TBATA* (B), *FAS* (C), *B3GAT1* (D) and *SLC52A3* (E)

Next, for each of the immune cell loci associated with both immune cell distributions and gene expression in blood we investigated whether the genetic variants responsible for both phenotypes were distinct or shared. To this end, for 20 of the 50 genome-wide significant immune cell type GWAS signals (**Table 1**), we performed colocalization analyses on a total of 47 genes (ranging from 1 to 14 genes per GWAS signal): These results yielded a posterior probability for a shared causal variant >80% (PP.H4>0.8) for half (10/20) of the GWAS signals. Four immune cell type GWAS signals showed evidence for colocalization with eQTL GWAS results for one gene each, while for the remaining six GWAS signals several colocalizations were observed (**Table 1, Supplementary Table 5**). Furthermore, we screened the colocalization results for suggestive signals (PP.H4>0.6) and identified two additional findings with *FAS* (PP.H4=0.79) for rs12257092 and *HNRNPLL* (PP.H4=0.64) for rs58905133.

### Association analyses of age, cytomegalovirus status, sex, and smoking

Linear regression analyses of the variables age, CMV status, sex, and smoking on the 92 immune cell types showed a substantial number of significant (FDR=0.01) associations: Specifically, age group, CMV status, and sex were significantly associated with 41 immune cell types (originating from 15 clusters), 36 immune cell types (11 clusters), and 13 immune cell types (five clusters), respectively (**Supplementary Table 10)**. Interestingly, smoking status was not found to show significant associations with any of the 92 analyzed phenotypes. Notably, CMV status was highly significantly associated (q-values <1.00E-14) with all CD4+ and CD8+ T cell subtypes in clusters 17 and 18 showing consistent (i.e. positive) directions of effect. Moreover, age was positively associated with the CD8+ memory cell subtypes in cluster 9 and with nearly all CD4+ T memory cells in cluster 10 and inversely associated with naïve, early and late differentiated CD8+ and CD4+ T cells in clusters 19 and 20. For sex, most of the significant associations were observed with different subsets of CD8+ cells.

### Genetic overlap with autoimmune diseases and COVID-19 using PGS analyses

Finally, we assessed whether genetic predisposition to five different autoimmune diseases and COVID-19 infection or long COVID manifests itself pre-disease by affecting the immune cell type composition of healthy adults. In these analyses, we observed 13 different significant associations showing permutation stability (p_empirical_<0.01) and passing FDR control at 1% for the T1D PRS: these predominately affected different subtypes of CD8+ cells with variances explained of 3-5% by the PRS (**Table 2**, **Supplementary Table 11**). Upon including only genome-wide significant T1D SNPs in the PRS, the vast majority (10/13) of the PRS analyses remained nominally significant significant (p<0.05), and one was borderline significant (**Supplementary Table 12**). Furthermore, we observed significant associations with early-differentiated CD4+ T cells for the long COVID PRS (variances explained: 3-4%; **Table 2**, **Supplementary Table 11**). None of the other PRS on additional autoimmune disease or other COVID-19 phenotypes showed significant associations here, although a few yielded nominally significant results that did not pass FDR and/or permutation control (**Supplementary Table 11)**. Finally, as expected, our height PRS analyses (negative control) did not show any significant results.

**Table 2.** Significant results for polygenic risk score analyses of immune cell distributions based on type 1 diabetes and long COVID risk GWAS. This table displays all significant results (q value<0.01) for the polygenic risk score (PRS) analyses of the immune cell distributions in BASE-II based on GWAS data for autoimmune disease risk and COVID-19 phenotypes. The GWAS on height was used as a negative control. The long COVID PRS listed here were based on GWAS summary statistics based on a strict case and control status (strict case definition = Long COVID after test-verified SARS-CoV2 infection, strict control definition = individuals that had SARS-CoV-2 but did not develop Long COVID). The type 1 diabetes GWAS was conducted by Chiou et al. Statistical significance was determined based on an empirical α=0.01 following 10,000 permutations and an FDR-adjusted p value (FDR=0.01). N=number, N PRS-SNPs=number of SNPs included in the best-performing PRS, threshold=p value threshold of the summary statistics for inclusion of SNPs in the PRS, PRS R2=increase of variance explained of immune cell phenotype by inclusion of PRS. Empirical p=p value after 10,000 rounds of permutation (assessing robustness of the result). q value=FDR-adjusted p value.

| Trait | N cases | N controls | N SNPs | Cluster | Short name | N PRS-SNPs | Threshold | PRS R2 | P | Empirical p | q value |
| --- | --- | --- | --- | --- | --- | --- | --- | --- | --- | --- | --- |
| Long COVID | 3,018 | 37,935 | 8,136,810 | 11 | CD4+ED_CD27-CD28+ | 15745 | 4.81E-02 | 0.0371 | 1.11E-04 | 2.80E-03 | 7.06E-03 |
|  |  |  |  | 17 | CD4+ED_CD27-CD28- | 15997 | 4.90E-02 | 0.0338 | 1.53E-04 | 3.40E-03 | 7.06E-03 |
| Type 1 diabetes | 18,942 | 501,638 | 9,074,688 | 8 | CD8+ME_CD57+ | 22692 | 4.13E-02 | 0.0274 | 6.73E-04 | 8.90E-03 | 4.97E-03 |
|  |  |  |  | 9 | CD8+CM_EM | 17170 | 2.69E-02 | 0.0299 | 3.18E-04 | 2.80E-03 | 3.25E-03 |
|  |  |  |  | 18 | CD8+CD27+ | 1014 | 2.00E-04 | 0.0390 | 4.33E-05 | 1.10E-03 | 9.95E-04 |
|  |  |  |  | 18 | CD8+CD28+ | 1196 | 3.00E-04 | 0.0368 | 6.89E-05 | 7.00E-04 | 1.06E-03 |
|  |  |  |  | 18 | CD8+CD57+ | 1988 | 8.50E-04 | 0.0361 | 6.31E-05 | 1.60E-03 | 1.06E-03 |
|  |  |  |  | 18 | CD8+TEMRA | 1014 | 2.00E-04 | 0.0329 | 2.11E-04 | 4.40E-03 | 2.72E-03 |
|  |  |  |  | 18 | CD8+LEM_CD57+ | 1014 | 2.00E-04 | 0.0324 | 2.37E-04 | 3.50E-03 | 2.72E-03 |
|  |  |  |  | 18 | CD8+LEM | 1112 | 2.50E-04 | 0.0306 | 3.69E-04 | 4.70E-03 | 3.39E-03 |
|  |  |  |  | 19 | CD4+LD | 26866 | 5.37E-02 | 0.0278 | 5.67E-04 | 7.30E-03 | 4.74E-03 |
|  |  |  |  | 20 | CD8+ED | 13679 | 1.91E-02 | 0.0492 | 2.87E-06 | 1.00E-04 | 2.10E-04 |
|  |  |  |  | 20 | CD8+LD_CD27+CD28+ | 13627 | 1.90E-02 | 0.0482 | 4.57E-06 | 1.00E-04 | 2.10E-04 |
|  |  |  |  | 20 | CD8+naïve | 13627 | 1.90E-02 | 0.0442 | 9.53E-06 | 2.00E-04 | 2.92E-04 |
|  |  |  |  | 20 | CD8+LD | 17170 | 2.69E-02 | 0.0271 | 7.02E-04 | 8.20E-03 | 4.97E-03 |
| Height | n.a. | 1,632,478 | 1,089,720 | - | - | - | - | - | - | - | - |

## Discussion

We performed a wide-ranging multi-omics study on the impact and possible functional role that genetic variants play in determining immune cell type composition in blood. In addition, we found that genetic predispositions to T1D and long COVID may affect immune cell composition in blood prior to clinical onset of the disease.

First, we completed genome-wide screenings on nearly 100 immune cell types where we validated the role of seven previously described loci and identified 13 novel loci that may impact the composition of immune cells in nearly 500 blood samples from immunologically healthy individuals. The most significant novel immune cell locus was *SLC52A3* that was associated with naïve CD57+ CD8+ T cells and showed colocalization with *SLC52A3* expression*. SLC52A3* encodes the “solute carrier family 52 member 3”, a riboflavin transporter protein that is strongly expressed in the intestine and plays a role in intestinal absorption of riboflavin (vitamin B2)^39^. Our GWAS now further supports a role of riboflavin in the distribution of naïve CD57+ CD8+ T cells in peripheral blood. Interestingly, the population of naïve CD57+ CD8+ T cells is only a very small subset of CD8+ cells (<1%) and has rarely been studied specifically. Thus, independent replication of this genome-wide significant finding will be required. In addition, we established novel genetic associations of immune system candidate regulators such as *TBATA* with the CD4+/CD8+ T cell ratio and *B3GAT1* with CD4+ and CD8+ early-memory T cells in peripheral blood. *TBATA* encodes “thymus, brain and testes associated”, a ligand for class I human leukocyte antigen in the thymus^40,41^ and *B3GAT1* encodes “beta-1,3-glucoronyltransferase 1”, a key enzyme in the biogenesis of HNK-1 (CD57) glycans, which has also been described to restrict influenza virus infection^42^. Notably, our extensive post-GWAS fine-mapping efforts showed that the majority of loci (including *SLC52A3, B3GAT1*, *FAS* and *HNRNPLL,* but not *TBATA*) showed robust *cis* effects on gene expression in blood and/or in relevant blood immune cells suggesting regulatory mechanisms on gene expression. This was in line with colocalization analyses with these QTL data that supported the role of *SLC52A3, B3GAT1*, *FAS* and *HNRNPLL* as immunological candidate genes, which should be considered for future genetic and functional studies. Interestingly, the composition of immune cells appears to be governed by both common (MAF>5%) and infrequent (MAF 1-5%) genetic variants.

In addition to considerably extending the list of candidate immune genes, our study also represents an important step forward in understanding the pathophysiological mechanisms underlying T1D and long COVID. This is based on the observation that the PRS of T1D predicts the proportion of CD8+ T cell subtypes in healthy adults. This is in line with previous reports that also suggest a central role of CD8+ cells in the onset of T1D (e.g., ref. ^43,44^). A second finding from this arm of the study was that early-differentiated CD4+ T cells are likely involved in predisposition to long COVID, which is in agreement with previous, functional data (e.g., ref. ^45,46^). Our novel results now show that the genetic predisposition to T1D and long COVID leads to different immune cell compositions of CD8+ and CD4+ T cells already in the pre-disease state.

The main strengths of our study are i) deep immune cell phenotyping, ii) a comparatively large sample size given the number of available immune cell phenotypes, iii) state-of-the-art computational analyses combining the main genomic findings with other omics (transcriptomics, epigenomics) domains. All of these led to the discovery of novel and immunologically plausible genetic associations between *SLC52A3, B3GAT1,* and *TBATA* and the distribution of immune cells in blood as well as genomic evidence supporting a central role of CD8+ T cells in the onset of T1D and of early-differentiated CD4+ T cells in long COVID. Despite these strengths, we note the following potential limitations of our work and results. First, while having examined a sample size of nearly 500 individuals in our GWAS, power was still limited to detect small effect sizes especially for infrequent SNPs. Thus, it is very likely that there remain many more genetic loci to be discovered with an impact on immune cell distributions in healthy adults once analyses on larger sample sizes are performed. Limited sample size may also be one of the reasons why we could replicate some, but not all, results for genetic loci described by previous GWAS on immune cell distributions. However, there are also other reasons for not replicating prior results such as (true) heterogeneity of effects across different populations, different sample ascertainment schemes and immune cell phenotyping protocols, different analysis designs, and type-1 errors in previous studies. Furthermore, despite using established thresholds of multiple testing correction, some of the novel immune loci highlighted in our study may nonetheless represent chance findings. Second, to the best of our knowledge this is the first study to examine the genetic overlap between immune cell distributions in blood of healthy adults and predisposition for autoimmune diseases and COVID-19 phenotypes. Despite leading to biologically highly plausible results, these findings need to be validated in independent datasets. Third, it is well known that the lead SNPs emerging from GWAS are not necessarily the variants exerting the molecular effects eliciting functional impacts. To this end, we used a range of state-of-the art fine-mapping strategies, including colocalization analyses, to pinpoint the functionally relevant gene(s) and potentially causal genetic variant(s). However, due to the large number of significant immune cell GWAS findings in this study and multiple QTL datasets available, we focused our analysis of gene expression data on statistically robust QTL effects in sufficiently sized samples (i.e., based on whole blood). This may have precluded the identification of more intricate molecular regulatory mechanisms on gene expression underlying the SNP-immune cell associations. Still, the colocalization analyses revealed several functionally meaningful findings such as *FAS*, *BGAT1*, and *HNRNPLL*. Finally, our dataset was comprised entirely of individuals of European-ancestry. Hence, we cannot make any inference on whether and which of the conclusions reached from our analyses are also applicable to individuals from other ethnic backgrounds.

In conclusion, our comprehensive multi-omics study identified genome-wide significant associations between thirteen novel loci and distinct immune cell types in peripheral blood and implicates riboflavin in the distribution of naïve CD57+ CD8+ T cells. Several of these associations appear to be founded gene expression effects. Importantly, the present study is the first to describe an association of genetic predisposition to T1D and long COVID with immune cell distribution in the blood of healthy adults, indicating a central role of CD8+ cells in the onset of T1D and of early-differentiated CD4+ T cells in long COVID. In addition to advancing our understanding of immune system function, these findings provide disease surrogates on the level of blood immune responses.

## Acknowledgements

We are grateful to all BASE-II participants. We thank Prof. An Goris for providing us with the GWAS summary statistics generated for the publication of Lagou et al., 2018^9^.

## Contributors

Study concept and supervision: CML, acquisition/contribution of data: DG, MV, JD, AG, ID, LB, GP, data analysis: LD, JH, DG, OO, SML, interpretation of results: LD, DG, FL, FZ, GP, LB, CML, Drafting the manuscript: LD, CML. Critical revision of the manuscript for content: all co-authors

## Data availability

Complete summary statistics of the GWAS analyses are publicly available at the GWAS Catalog (https://www.ebi.ac.uk/gwas/home, accession number: *pending*) as well as at the Zenodo platform (https://zenodo.org, study data accessible at: *pending*). Subject-level data can be obtained by qualified investigators upon request to the authors.

## References

1. Theofilopoulos, A. N., Kono, D. H. & Baccala, R. The multiple pathways to autoimmunity. Nat. Immunol. 18, 716–724 (2017).

2. Leone, R. D. & Powell, J. D. Metabolism of immune cells in cancer. Nat. Rev. Cancer 20, 516–531 (2020).

3. Patin, E. et al. Natural variation in the parameters of innate immune cells is preferentially driven by genetic factors. Nat. Immunol. 19, (2018).

4. Bossini-Castillo, L. et al. Genomic Risk Score impact on susceptibility to systemic sclerosis. Ann. Rheum. Dis. 80, 118–127 (2021).

5. Gutierrez-Arcelus, M., Rich, S. S. & Raychaudhuri, S. Autoimmune diseases — connecting risk alleles with molecular traits of the immune system. Nat. Rev. Genet. 17, 160–174 (2016).

6. Orrù, V., Steri, M., Cucca, F. & Fiorillo, E. Application of Genetic Studies to Flow Cytometry Data and Its Impact on Therapeutic Intervention for Autoimmune Disease. Front. Immunol. 12, (2021).

7. Orrù, V. et al. Complex genetic signatures in immune cells underlie autoimmunity and inform therapy. Nat. Genet. 52, 1036–1045 (2020).

8. Roederer, M. et al. The Genetic Architecture of the Human Immune System: A Bioresource for Autoimmunity and Disease Pathogenesis. Cell 161, 387–403 (2015).

9. Lagou, V. et al. Genetic Architecture of Adaptive Immune System Identifies Key Immune Regulators. Cell Rep. 25, 798–810.e6 (2018).

10. Aguirre-Gamboa, R. et al. Differential Effects of Environmental and Genetic Factors on T and B Cell Immune Traits. Cell Rep. 17, 2474–2487 (2016).

11. Orrù, V. et al. Genetic Variants Regulating Immune Cell Levels in Health and Disease. Cell 155, 242–256 (2013).

12. Brodin, P. et al. Variation in the Human Immune System Is Largely Driven by Non-Heritable Influences. Cell 160, 37–47 (2015).

13. Bertram, L. et al. Cohort Profile: The Berlin Aging Study II (BASE-II). Int. J. Epidemiol. 43, 703–712 (2014).

14. Lill, C. M. et al. Genetic Burden Analyses of Phenotypes Relevant to Aging in the Berlin Aging Study II (BASE-II). Gerontology 62, 316–322 (2016).

15. Sbierski-Kind, J. et al. T cell phenotypes associated with insulin resistance: results from the Berlin Aging Study II. Immun. Ageing 17, (2020).

16. Deecke, L., et al. No increase of CD8+ TEMRA cells in the blood of healthy adults at high genetic risk of Alzheimer’s disease. Alzheimer’s Dement. J. Alzheimer’s Assoc. [in Press.(2023).

17. Charrad, M., Ghazzali, N., Boiteau, V. & Niknafs, A. NbClustlll: An R Package for Determining the Relevant Number of Clusters in a Data Set. J. Stat. Softw. 61, (2014).

18. Chang, C. C. et al. Second-generation PLINK: rising to the challenge of larger and richer datasets. Gigascience 4, (2015).

19. Hong, S. et al. Genome-wide association study of Alzheimer’s disease CSF biomarkers in the EMIF-AD Multimodal Biomarker Discovery dataset. Transl. Psychiatry 10, (2020).

20. Devlin, B. & Roeder, K. Genomic Control for Association Studies. Biometrics 55, 997–1004 (1999).

21. Yin, L. et al. rMVP: A Memory-efficient, Visualization-enhanced, and Parallel-accelerated tool for Genome-wide Association Study. Genomics. Proteomics Bioinformatics (2021) doi:10.1016/j.gpb.2020.10.007.

22. Kircher, M. et al. A general framework for estimating the relative pathogenicity of human genetic variants. Nat. Genet. 46, (2014).

23. Plagnol, V., Smyth, D. J., Todd, J. A. & Clayton, D. G. Statistical independence of the colocalized association signals for type 1 diabetes and RPS26 gene expression on chromosome 12q13. Biostatistics 10, 327–34 (2009).

24. Zheng, Z. et al. QTLbase: an integrative resource for quantitative trait loci across multiple human molecular phenotypes. Nucleic Acids Res. 48, D983–D991 (2020).

25. Võsa, U. et al. Large-scale cis- and trans-eQTL analyses identify thousands of genetic loci and polygenic scores that regulate blood gene expression. Nat. Genet. 53, 1300–1310 (2021).

26. Giambartolomei, C. et al. Bayesian Test for Colocalisation between Pairs of Genetic Association Studies Using Summary Statistics. PLoS Genet. 10, e1004383 (2014).

27. Buniello, A. et al. The NHGRI-EBI GWAS Catalog of published genome-wide association studies, targeted arrays and summary statistics 2019. Nucleic Acids Res. 47, (2019).

28. Choi, S. W. & O’Reilly, P. F. PRSice-2: Polygenic Risk Score software for biobank-scale data. Gigascience 8, (2019).

29. Patsopoulos, N. A. et al. Multiple sclerosis genomic map implicates peripheral immune cells and microglia in susceptibility. Science (80-.). 365, (2019).

30. Chiou, J. et al. Interpreting type 1 diabetes risk with genetics and single-cell epigenomics. Nature 594, 398–402 (2021).

31. Ha, E., Bae, S.-C. & Kim, K. Large-scale meta-analysis across East Asian and European populations updated genetic architecture and variant-driven biology of rheumatoid arthritis, identifying 11 novel susceptibility loci. Ann. Rheum. Dis. 80, 558–565 (2021).

32. de Lange, K. M. et al. Genome-wide association study implicates immune activation of multiple integrin genes in inflammatory bowel disease. Nat. Genet. 49, 256–261 (2017).

33. COVID-19 Host Genetics Initiative. Mapping the human genetic architecture of COVID-19. Nature 600, 472–477 (2021).

34. Lammi, V. et al. Genome-wide Association Study of Long COVID. medRxiv 2023.06.29.23292056 (2023) doi:10.1101/2023.06.29.23292056.

35. Yengo, L. et al. A saturated map of common genetic variants associated with human height. Nature 610, 704–712 (2022).

36. Appay, V., van Lier, R. A. W., Sallusto, F. & Roederer, M. Phenotype and function of human T lymphocyte subsets: consensus and issues. Cytometry. A 73, 975–83 (2008).

37. Seder, R. A. & Ahmed, R. Similarities and differences in CD4+ and CD8+ effector and memory T cell generation. Nat. Immunol. 4, 835–842 (2003).

38. Kent, W. J. et al. The Human Genome Browser at UCSC. Genome Res. 12, 996–1006 (2002).

39. Brown, G. R. et al. Gene: a gene-centered information resource at NCBI. Nucleic Acids Res. 43, D36–D42 (2015).

40. Espinosa, G. et al. Peptides presented by HLA class I molecules in the human thymus. J. Proteomics 94, 23–36 (2013).

41. Flomerfelt, F. A. et al. Tbata modulates thymic stromal cell proliferation and thymus function. J. Exp. Med. 207, 2521–2532 (2010).

42. Trimarco, J. D. et al. Cellular glycan modification by B3GAT1 broadly restricts influenza virus infection. Nat. Commun. 13, 6456 (2022).

43. Gearty, S. V. et al. An autoimmune stem-like CD8 T cell population drives type 1 diabetes. Nature 602, 156–161 (2022).

44. Shimokawa, C. et al. CD8+ regulatory T cells are critical in prevention of autoimmune-mediated diabetes. Nat. Commun. 11, 1922 (2020).

45. Klein, J. et al. Distinguishing features of long COVID identified through immune profiling. Nature 623, 139–148 (2023).

46. Su, Y. et al. Multiple early factors anticipate post-acute COVID-19 sequelae. Cell 185, 881–895.e20 (2022).

47. Wang, K., Li, M. & Hakonarson, H. ANNOVAR: functional annotation of genetic variants from high-throughput sequencing data. Nucleic Acids Res. 38, e164–e164 (2010).

