## Supplementary Material for "Genome-wide association study reveals different T cell distributions in peripheral blood of healthy individuals at high genetic risk of type 1 diabetes and long COVID"

**Supplementary Methods**

*Study participants:* In this study, we investigated an effective sample size of 483 healthy participants of European descent from the Berlin Aging Study II (BASE-II) in Germany, a multi-institutional and multi-disciplinary longitudinal study aimed at investigating factors modulating the aging process^1,2^. All participants were part of the baseline recruitment of BASE-II and comprised a group of 344 older (aged 60-82 years) and 139 younger adults (aged 23-35 years, **Supplementary Table 2)** for whom quality-controlled genetic and phenotypic data derived from peripheral blood mononuclear cells (PBMC) were available. As the aim of the study was to assess determinants of the immune system in healthy individuals, we only included “immunologically healthy” BASE-II participants, defined as being without prevalent immune system-related diseases or treatment at baseline, i.e., autoimmune diseases (rheumatoid arthritis (RA), psoriatic arthritis, systemic lupus erythematosus, type 1 diabetes (T1D), multiple sclerosis (MS), Crohn's disease (CD), ulcerative colitis (UC), psoriasis, celiac disease, lymphocytic colitis), HIV, acute fever (defined as current or within the previous six weeks), receiving immunomodulatory systemic therapy, or with CRP values >10mg/L in plasma drawn at baseline.

*Generation of immune cell data by flow cytometry:* The isolation of PBMCs from whole blood samples of BASE-II participants and subsequent flow cytometry was performed at the Department of Immunology, University of Tübingen, Germany, as previously described in refs. ^3,4^. Briefly, PBMCs were isolated by density gradient centrifugation, cryopreserved and biobanked in liquid nitrogen. For the immunological analysis, PBMCs were thawed and two different antibody panels for flow cytometry panels were applied, each quantifying functionally related immune cell subsets^3^. Panel 1 encompassed a total of 50 immune cell types including various differentiated T lymphocyte phenotypes, and panel 2 quantified 41 immune cell types including monocytes, natural killer (NK) cells, NKT cells, myeloid-derived suppressor cells (MDSC), B cells, and general T cell populations (see **Supplementary Figure 1**, **Supplementary Table 3** for a detailed listing of all investigated immune cell types). For flow cytometry, after thawing, PBMCs were incubated with GAMUNEX (human IgG from Bayer, Leverkusen, Germany) and ethidium monoazide (EMA) bromide (MoBiTec GmbH, Göttingen, Germany) to reduce nonspecific binding of antibodies through Fc receptor blockade and to exclude EMA+ dead cells from the analysis. For panel 1, cells were then incubated with anti CCR7 (CD197) (R&D Systems, [Minneapolis](https://www.google.com/search?client=firefox-b-d&channel=trow5&q=Minneapolis&stick=H4sIAAAAAAAAAOPgE-LVT9c3NExKNk02rMiqUOLUz9U3SCuoKk_T0spOttLPL0pPzMusSizJzM9D4VhlpCamFJYmFpWkFhUvYuX2zczLS00syM_JLN7ByggA7JP4C1kAAAA&sa=X&ved=2ahUKEwjFs4bah8r2AhWFuKQKHYtQDVsQmxMoAXoECBwQAw), Minnesota, USA), anti-mouse IgG Pacific Orange (Invitrogen, [Waltham, Massachusetts,](https://www.google.com/search?client=firefox-b-d&channel=trow5&q=Waltham&stick=H4sIAAAAAAAAAOPgE-LUz9U3MDNLKUxS4gAxM6qMTbW0spOt9POL0hPzMqsSSzLz81A4VhmpiSmFpYlFJalFxYtY2cMTc0oyEnN3sDICANGzN1FQAAAA&sa=X&ved=2ahUKEwiElIr4h8r2AhUsPewKHZRfA2oQmxMoAXoECBcQAw) USA) and mouse serum. Next, for both panels, cells were stained for 20 min on ice with monoclonal antibodies against the markers listed in **Supplementary Table 4.** The cells were acquired with a 3 laser BD LSRII (BD biosciences, Heidelberg, Germany) flow cytometer and DIVA6 software. Samples were analyzed with FlowJo version 7.5 (TreeStar, Portland, USA). To assure constant cytometer performance, BD CST beads (BD biosciences) were run on each measurement day and as an additional reference control, PBMCs from the same biobanked donor were analyzed on each day of the quantifications. To set up adequate gates for analysis, fluorescence minus one controls were included in this study. Where applicable, following visual inspection, phenotypic values were transformed using log, root, log(100-x) and square transformations, respectively (**Supplementary Figure 2,** **Supplementary Table 3**). After transformation, outlier values were determined (defined as values <Q1-1.5*IQR or >Q3+1.5*IQR) and excluded from the subsequent analyses. Following the transformation, we excluded BASE-II participants with missing or outlier values across more than one third of all immune cell type data points resulting in the exclusion of 11 individuals for panel 1 and one individual for panel 2. Following these and additional QC steps, the effective sample size included in all subsequent statistical analyses was 483 (420 individuals for panel 1 and 352 individuals for panel 2; **Supplementary Table 3** for exact sample sizes per GWAS).

Some of the immune cell types analyzed in this study correlated with each other. Thus, we performed hierarchical clustering (calculated with R package NbClust^5^, [ward method, Euclidean distance]) to assess the interdependencies of the immune cell types. Accordingly, where appropriate, we present the GWAS results by clusters (e.g., in the “Candy plot” mentioned below). The optimal number of clusters that best represents the data structure was determined by the majority rule on a combination of cluster analysis methods implemented in NbClust.

*Generation of genome-wide SNP data:* The generation of genome-wide SNPdata, QC, and imputation has been performed as previously described^4,6^ : DNA for the larger BASE-II cohort (~2600 BASE-II participants) was extracted from whole-blood EDTA samples. Genome-wide microarray-based genotyping on the DNA was performed using the Genome-Wide Human SNP Array 6.0 (Affymetrix Inc., Santa Clara, California, USA) (e.g., ref. ^2^). All data processing and analysis steps were performed with PLINK v1.9 (www.cog-genomics.org/plink/1.9/) or v2.0 (www.cog-genomics.org/plink/2.0/)^7^, unless stated otherwise. Prior to imputation, the genotype data underwent extensive QC: Briefly, sample-based pre-imputation QC excluded individuals with mismatches of genetic vs reported sex (--check-sex 0.25 0.75), individuals with high missing genotype rate (--mind 0.05) and individuals with extreme heterozygosity (--het; mean heterozygosity of the population ±6 standard deviations). After linkage disequilibrium (LD)-pruning (--indep-pairwise 1500 150 0.2), pairwise allele-sharing identity-by-descent (IBD)/identity-by-state was determined using (--Z-genome --min 0.1), and individuals exceeding an IBD threshold of 0.1 were excluded. The LD-pruned dataset was also used for principal component analysis (PCA; --pca) using the 1000 Genomes Project Consortium Phase 3 dataset^8^ as reference for the classification of individuals of non-European ethnicity (controlling the false discovery rate [FDR] at 5%). SNP-based pre-imputation QC included the following steps: strand check (--flip), missing genotype rate (--geno 0.02), Hardy-Weinberg equilibrium (HWE) tests (--hwe 0.000001; based on Fisher’s^9^ exact test), and minor allele frequency (MAF) filtering (--maf 0.01). Bcftools (v.1.9) was used to remove ambiguous SNPs and, where applicable, to swap alleles to match the human reference genome GRCh37/hg19. After QC, 723,727 genotyped SNPs were available in 2,351 subjects. Next, we performed genotype imputation using the Haplotype Reference Consortium (HRC) reference panel and Minimac3 as described previously^4,6^ resulting in a total of 38,407,851 SNPs. For post-imputation QC, the same pre-imputation QC steps were applied (see above). In addition, we excluded newly imputed SNPs with an imputation quality score R^2^<0.7. Furthermore, for SNPs showing evidence for association at a *p*<1.00E-5 in the subsequent GWAS (see below), we compared the MAF observed in BASE-II with that reported in the 1000 Genomes dataset (EUR population) and excluded SNPs from our report that differed by more than 10% in the MAF between both datasets, resulting in the exclusion of eight SNPs. Thus, the final QC’ed dataset comprised 2,342 BASE-II participants and 6,932,885 genotyped and imputed SNPs with MAF ≥0.01. Merging the effective sample size from the immunological assays with the SNP genotyping data resulted in 483 participants (420 individuals for panel 1 and 352 individuals for panel 2; **Supplementary Table 3** for exact sample sizes per GWAS analysis) and 6,932,885 SNPs.

*Statistical analyses on immune cell distributions:* Following LD pruning, we performed PCA on each of the four effective datasets (i.e., for the datasets on the two immune cell panels stratified for older and younger BASE-II participants). GWAS were performed on 51 immune cell phenotypes from panel 1 and 41 phenotypes from panel 2 based on linear regression analyses adjusting for sex and PCs 1-4 using PLINK v2.0^7^. GWAS for each immune cell subpopulation were run separately for both age groups and combined by fixed-effect meta-analyses using PLINK v1.9^7^. The heterogeneity of the effects estimated in the two groups was quantified using the $I^{2}$ statistic. Possible inflation of the test statistics was assessed by visual inspection of the corresponding quantile-quantile plots (QQ plots, generated using the Functional Mapping and Annotation (FUMA) platform^10^ (<https://fuma.ctglab.nl>)) and by calculation of the respective inflation factor λ^11^. We visualized our main results in a Manhattan plot generated by the R package CMplot^12^, which allowed us to display all GWAS results in one plot by color-coding the different cell type clusters (“Candy plot”). Genome-wide significance was defined at an α=5.00E-8. Genome-wide significantly associated index SNPs were defined as the most significantly associated SNP per locus (i.e., ±1Mb around the most significantly associated SNP). For the genome-wide significantly associated region on chromosome 1 (*MIR181A1HG/PTPRC* locus), the window defining a locus was extended to ±2Mb based on an extended LD in this region. To identify additional independent signals for each locus with a genome-wide significant association signal, we performed linear regression analyses conditioning on the index SNP in a ±1Mb window using PLINK2. Furthermore, we performed sensitivity analyses by performing regression analyses conditioning on the most significant index SNP on chromosome 8q21.11 (rs117904683) in an extended window of ±3Mb (to verify the independent nature of the two index SNPs in that region). The conditional results were FDR-controlled at 1%. In addition, we examined the previously reported associations of age (older vs younger age group), CMV status (positive vs negative), sex (women vs men), and smoking (current vs non-current [former/never smokers]), with the immune cell phenotypes using multivariate linear regression (lm function) in R. The results were FDR-controlled at 1%.

*Functional annotations of GWAS results:* Index SNPs and their proxies (r^2^>0.6) were annotated according to genomic location, nearest gene, functional consequence, CADD score^13^, molecular *cis* quantitative trait loci (QTL) effects, and colocalization analysis results^14^. *Cis* QTL effects were assessed using the QTLbase^15^ (<http://www.mulinlab.org/qtlbase/index.html>, downloaded on March 16th, 2022). Specifically, we considered *cis* QTL results for 4 molecular markers (i.e., expression QTL [eQTL], protein QTL [pQTL], methylation QTL [mQTL] and histone modification QTL [hQTL]) in whole blood and in a selection of 12 blood cell types depending on their relevance to the respective GWAS finding (i.e., B cells, monocytes, CD14+ monocytes, NK cells, CD16+ neutrophils, CD4+ T cells, naive CD4+ T cells, CD8+ T cells, activated CD8+ T cells, naive CD8+ T cells, and lymphocytes). In this context, the type-1 error rate for the QTL effect of an index SNP was set to α=1.00E-5. We assessed a shared genetic etiology for both the immune cell GWAS signal and gene expression signals in the same locus by performing colocalization analyses^14^ of each GWAS region (index SNP ±1Mb) using the largest blood eQTL dataset published to date^16^ (n=31,684; data downloaded from QTLbase on May 04^th^, 2022) based on the R package *coloc* v5 (<https://cran.r-project.org/web/packages/coloc/index.html>)^17^ (**Table 1, Supplementary Table 5**). We limited the investigated QTL SNPs to those showing significant eQTL effects with p<1.00E-5. No further pruning or trimming was applied. Colocalization was defined as the posterior probability of a shared causal variant >80% (‘PP.H4’>0.8). In addition, we scanned the colocalization results for suggestive colocalized hits by relaxing the threshold to PP.H4> 0.6. Furthermore, we compared our genome-wide significant findings with those from previous immune cell GWAS^18–22^ by using the data provided in the original publication^18–21^ or by using GWAS summary statistics made available to us by the investigators^22^ (**Table 1, Supplementary Table 1**), and to immunologically relevant phenotypes listed in the GWAS Catalog^23^.

*Polygenic risk score analyses:* Next, we assessed whether autoimmune disease or COVID-19 predisposition already manifests itself by different immune cell type compositions in immunological healthy adults. For this, we calculated best-fitting PRS for autoimmune diseases and COVID-19 based on publicly available GWAS summary statistics: For autoimmune diseases we used available GWAS summary statistics on MS^24^ , T1D^25^ , RA^26^, CD^27^ and UC^27^. For COVID-19, we used summary statistics from three COVID-19 case-control GWAS datasets^28^ (<https://www.covid19hg.org/results/r5/>) and from a recent GWAS on long COVID^29^ (comprising four largely similar datasets using strict or broader case/control definitions, e.g. https://my.locuszoom.org/gwas/793752/?token=0dc986619af14b6e8a564c580d3220b4). As a negative control, we used summary statistics from a large GWAS dataset on human height (ref. ^30^). PRS were calculated using the default parameters of PRSice-2^31^. Prior to PRS generation, ambiguous SNPs, duplicate entries, and SNPs with MAF <5% were excluded from the GWAS datasets. Next, we performed linear regression analyses on the PRS and the 92 immune cell types in the BASE-II dataset adjusting for sex and the first four PCs using PRSice-2. Empirical p values were calculated using 10,000 permutations (--perm 10000). We chose a conservative approach and defined a result as being statistically significant if it showed empirical significance (α_emp_=0.01) following permutation and if it passed FDR control at 1%.

**Supplementary Figure 1. Overview of the immune cell types assessed in the current study**

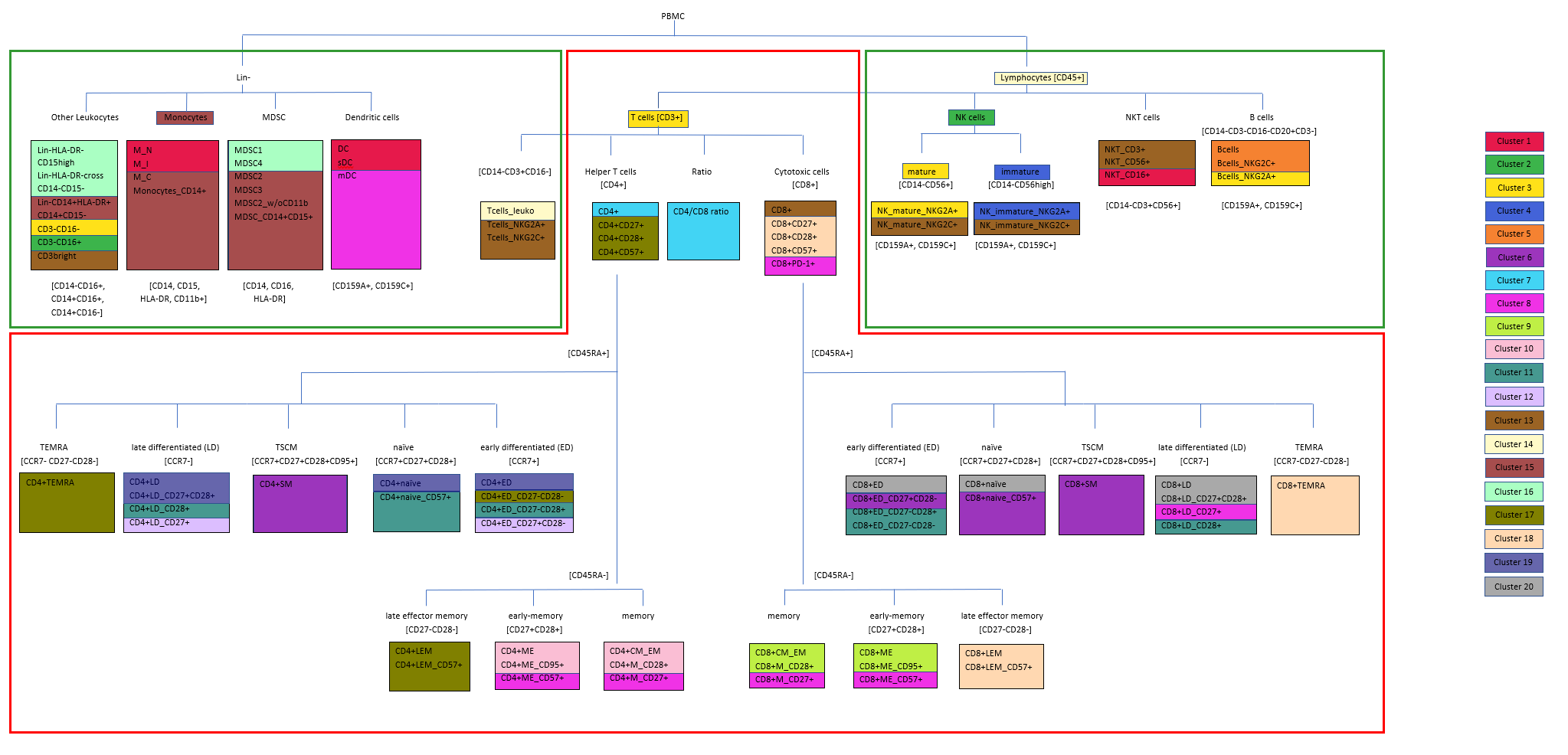

Legend. This diagram displays all immune cell phenotypes analyzed in this study. The cell types are colored according to the cluster they belong to (see **Supplementary Table 3**). All cell types framed by the red box belong to panel 1 and those framed by the green box belong to panel 2 of the flow cytometry. PBMC = peripheral blood mononuclear cell; Lin- = lineage-negative cells (no B, NK or T cells); MDSC = myeloid derived suppressor cells; NK cells = natural killer cells; NKT cells = natural killer T cells; TEMRA = effector memory cells re-expressing CD45RA; TSCM = T-stem cell-like memory

**Supplementary Figure 2:** Immune cell type distributions in BASE-II before and after transformation and outlier removal

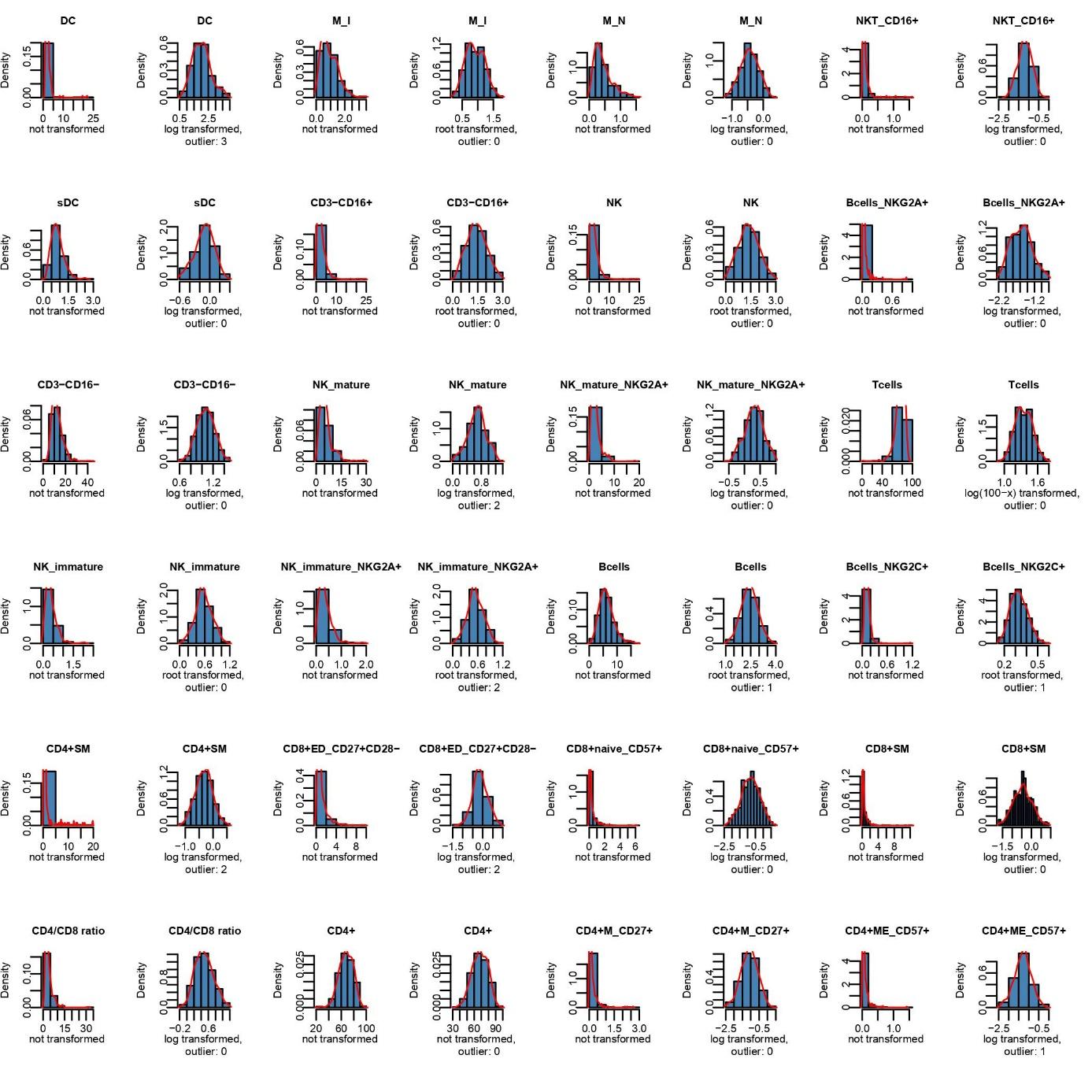

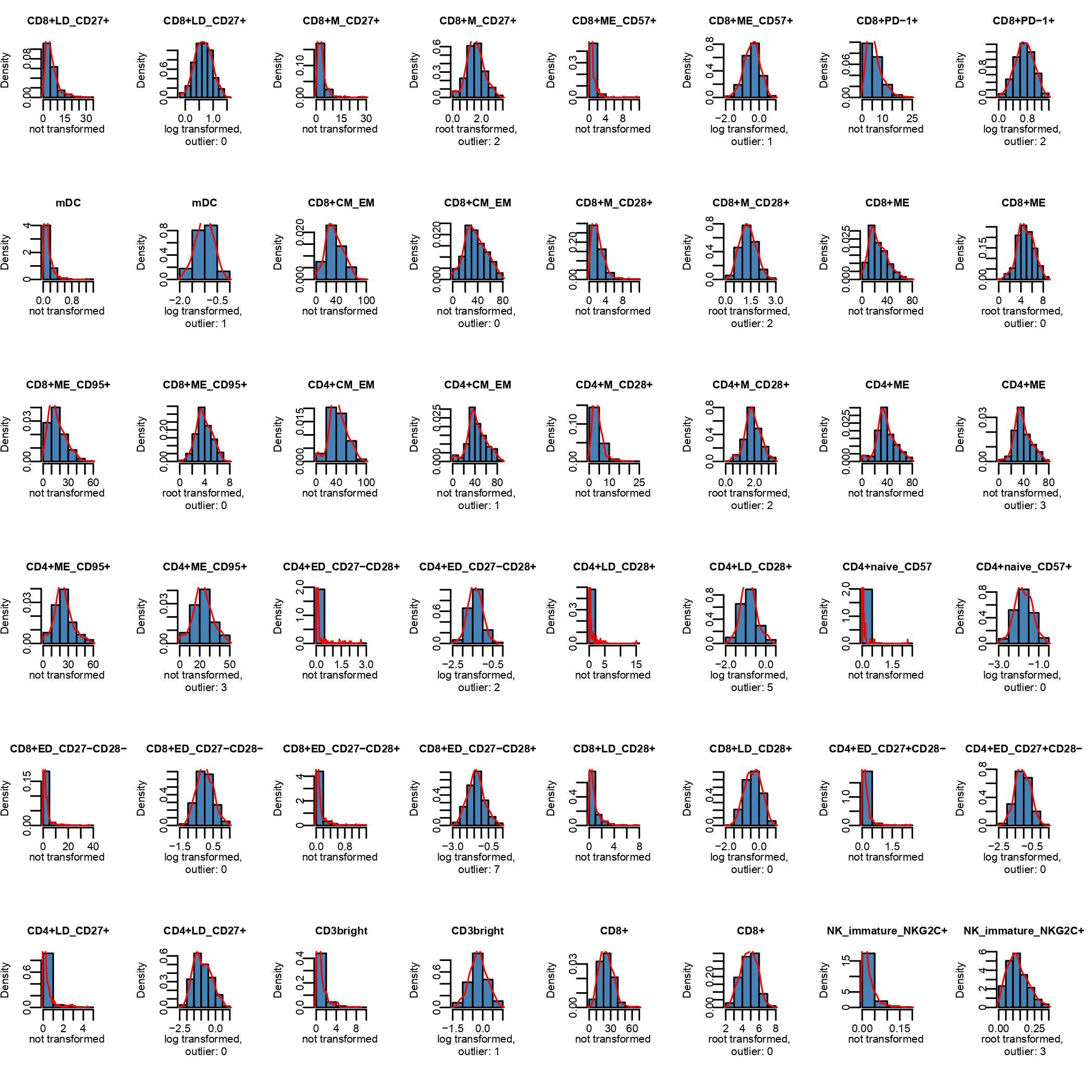

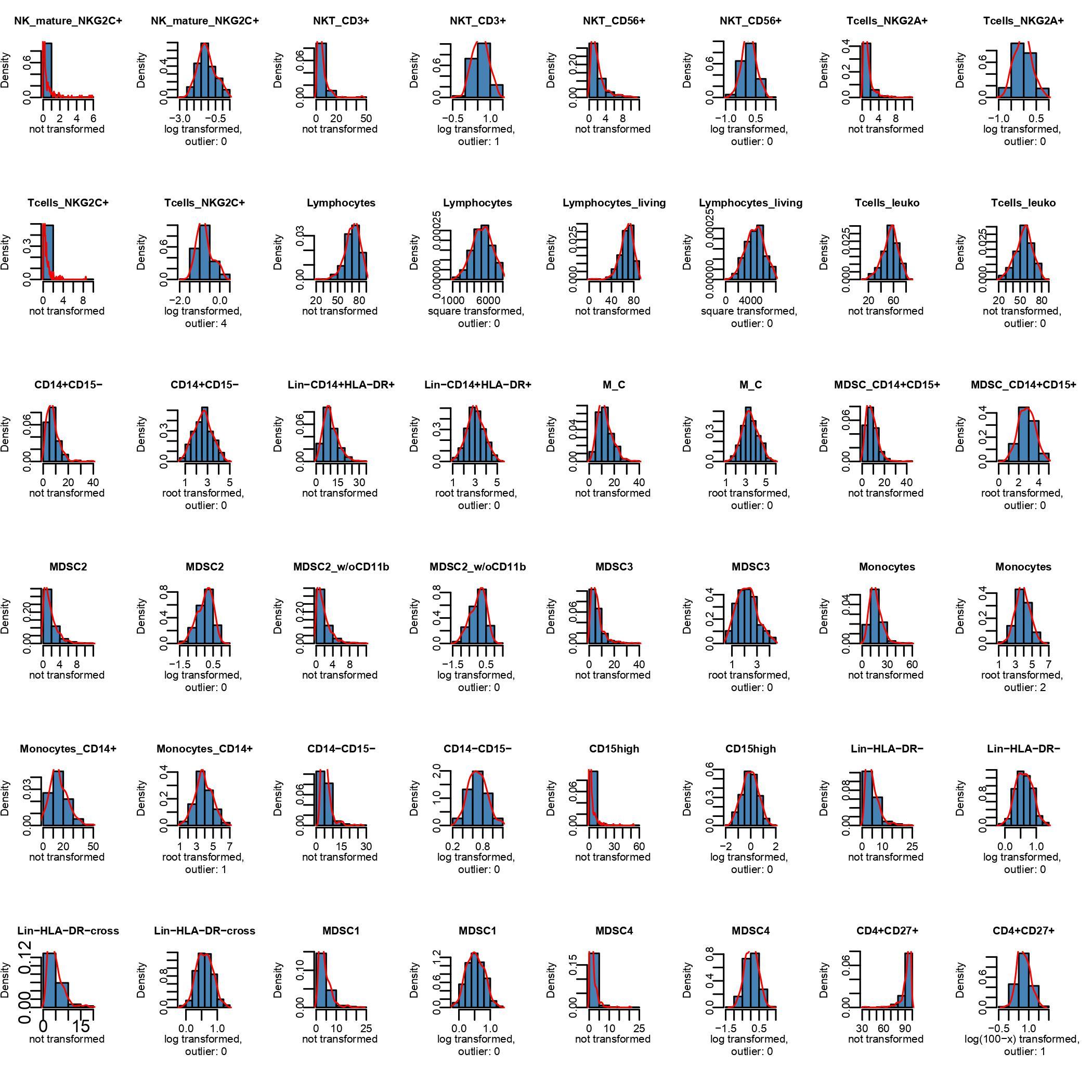

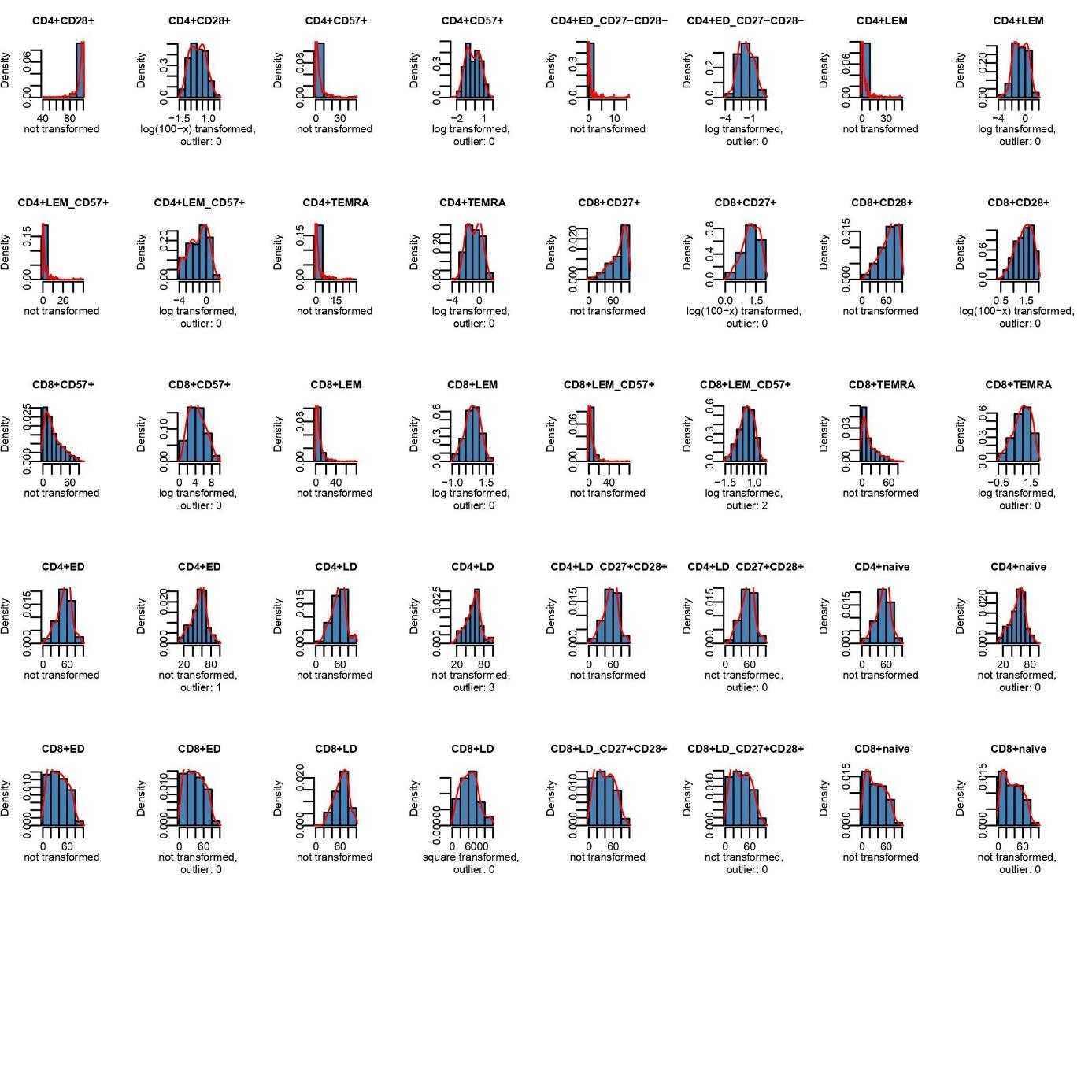

Legend. Data were transformed, where applicable, using log, root, log(100-x) or square transformations. For each phenotype the untransformed data are displayed followed by the transformed data where appropriate. The number of outliers which were removed after transformation is also displayed (see Methods section for more details).

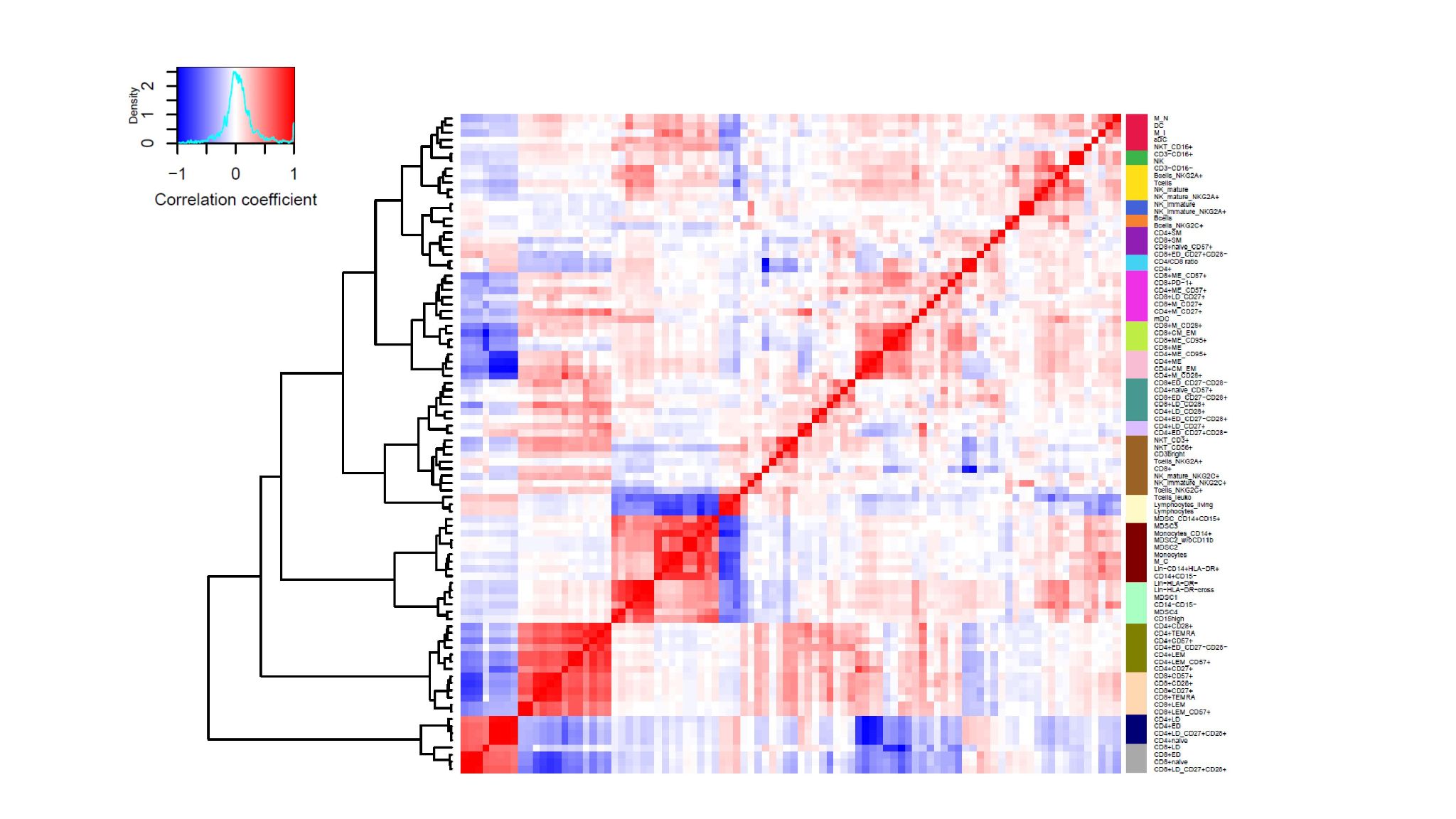
**Supplementary Figure 3:** Heatmap of correlations of the immune cell phenotypes quantified in BASE-II

Legend. The clusters were calculated with the ward hierarchical clustering method of the NbClust R package^5^ based on Spearman’s correlation coefficient.

**Supplementary Figure 4:** Quantile-quantile plots of *p* values of immune cell GWAS in BASE-II

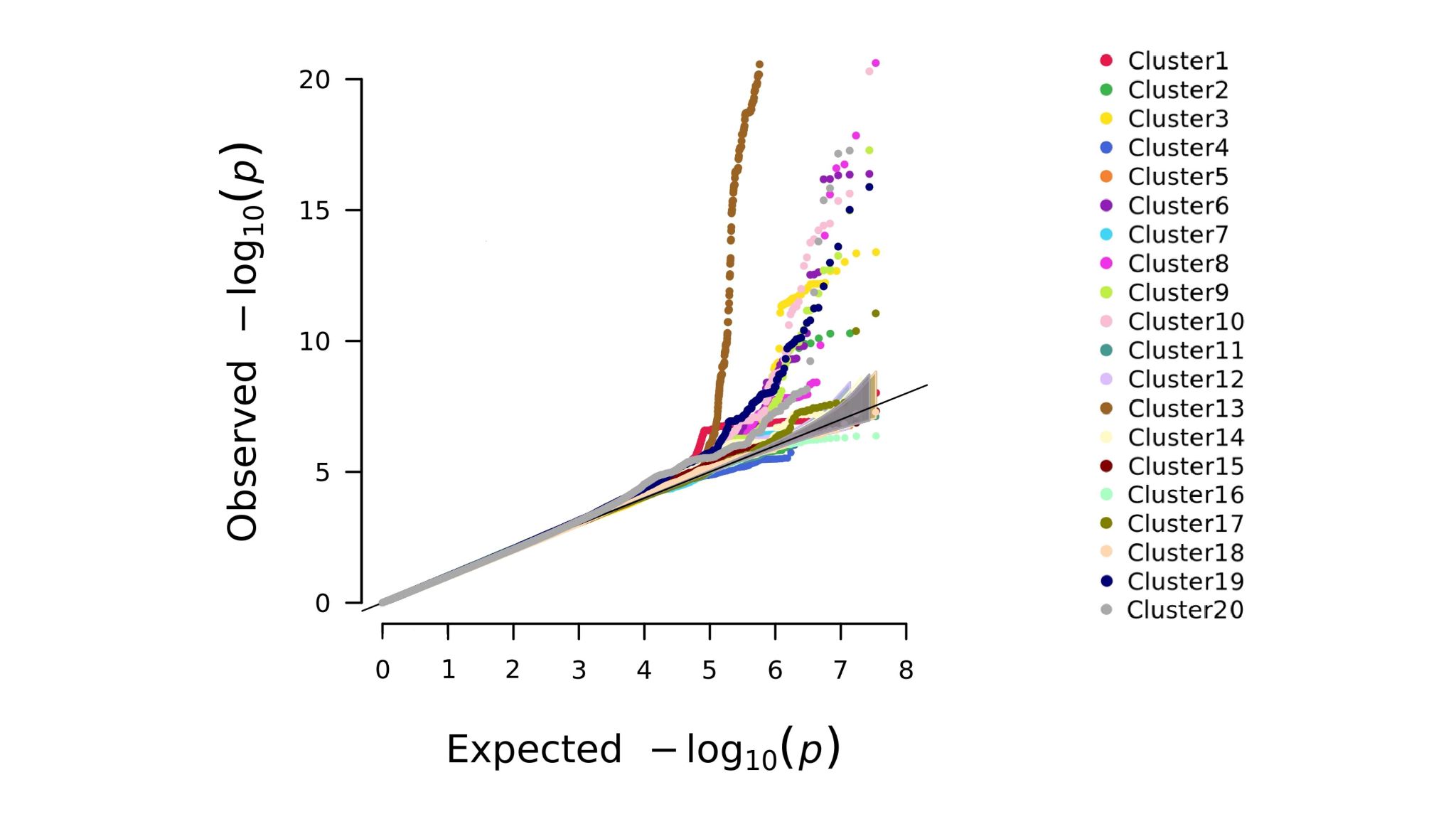
 A.

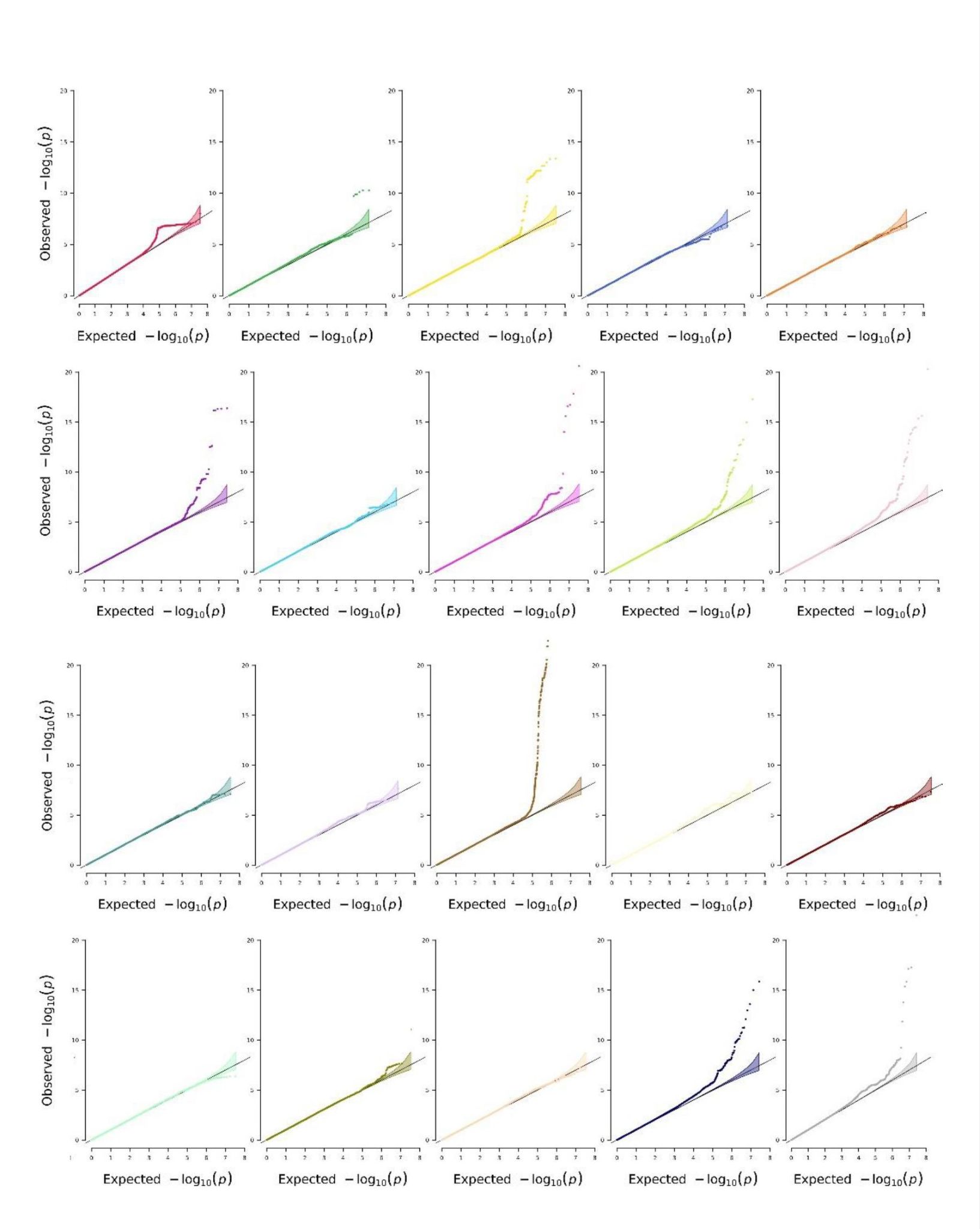

B.

Legend. This figure displays the quantile-quantile (QQ) plots for all immune cell GWAS. The QQ plots were subsumed in 20 clusters and displayed in a single QQ plot (A) as well as for each cluster separately (B). The inflation factor λ showed no substantial inflation for the individual immune cell GWAS (range of λ = 0.99-1.03).

**Supplementary Figure 5:** Overview of association results of genome-wide significant SNPs for all immune cell phenotypes

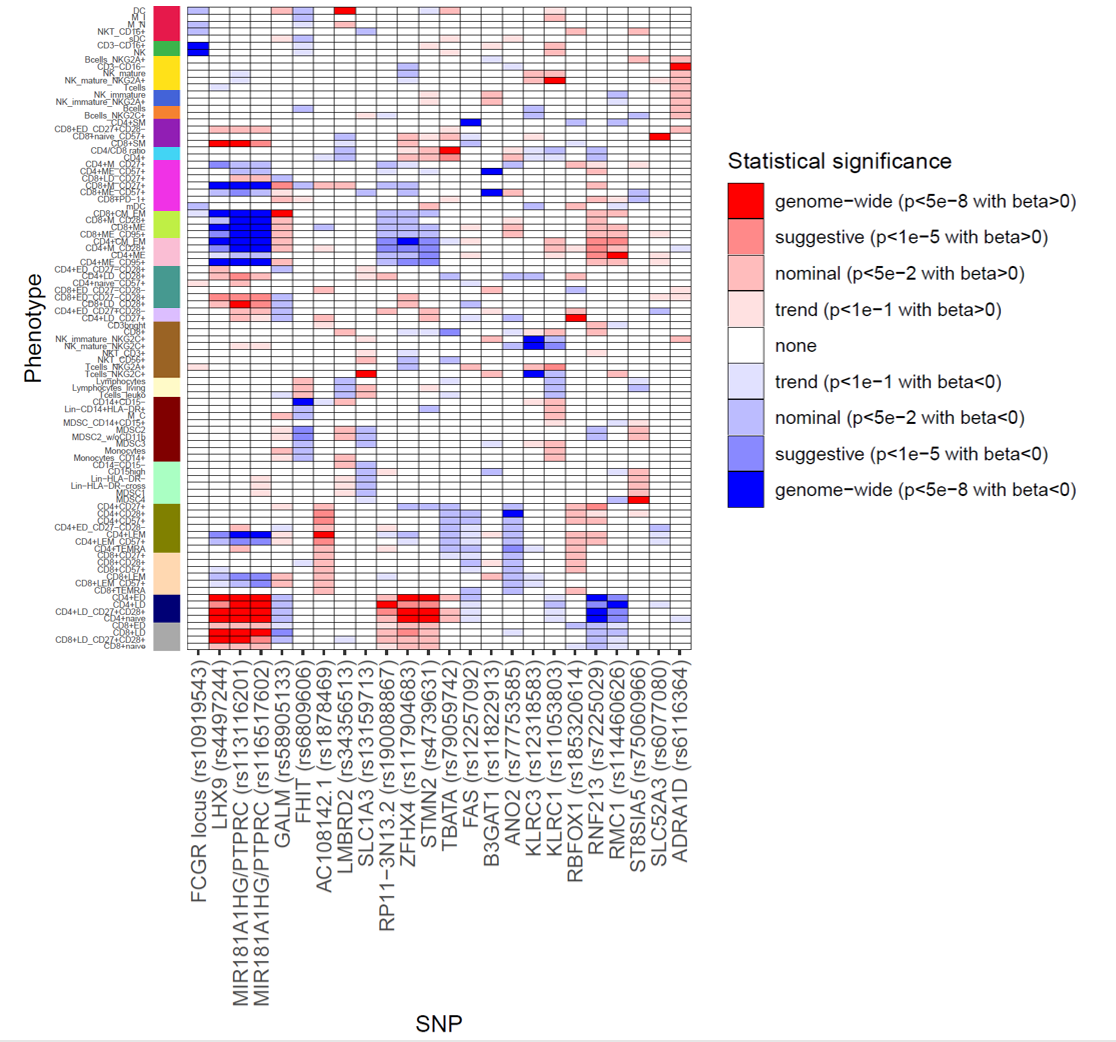

Legend. This figure visualizes the association results of all SNPs that are genome-wide significantly (α=5.00E-8) associated with at least one immune cell type in BASE-II in all immune cell types. Red indicates positive, blue indicates negative betas with respect to the minor allele. The leftmost column denotes the different immune cell type clusters.

**Supplementary Figure 6:** Locus zoom plots for the 24 genome-wide significant index SNPs

A. rs10919543 (*FCGR* locus; NK)

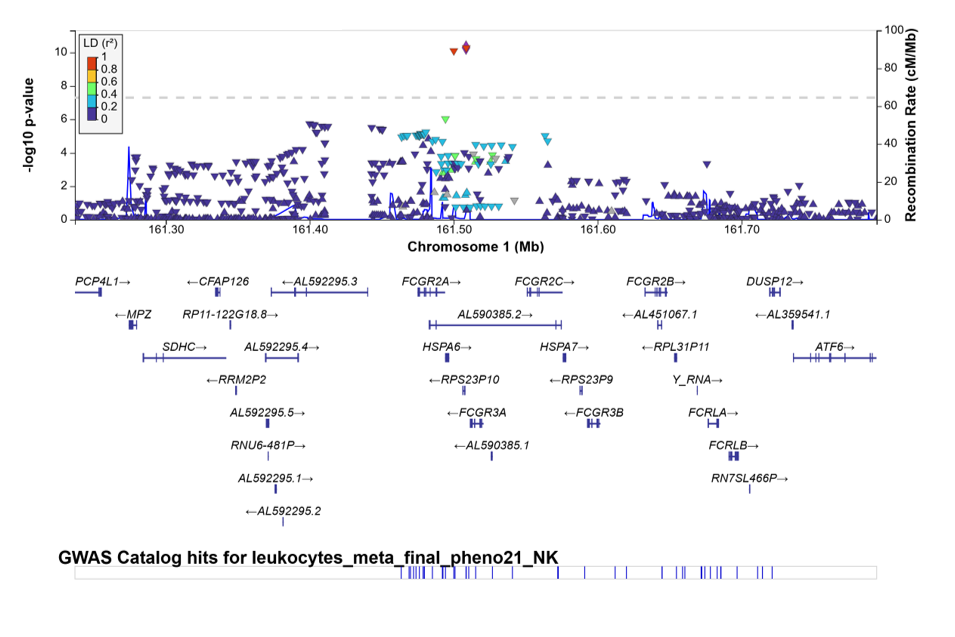

B. rs4497244 (*LHX9*; CD8+SM)

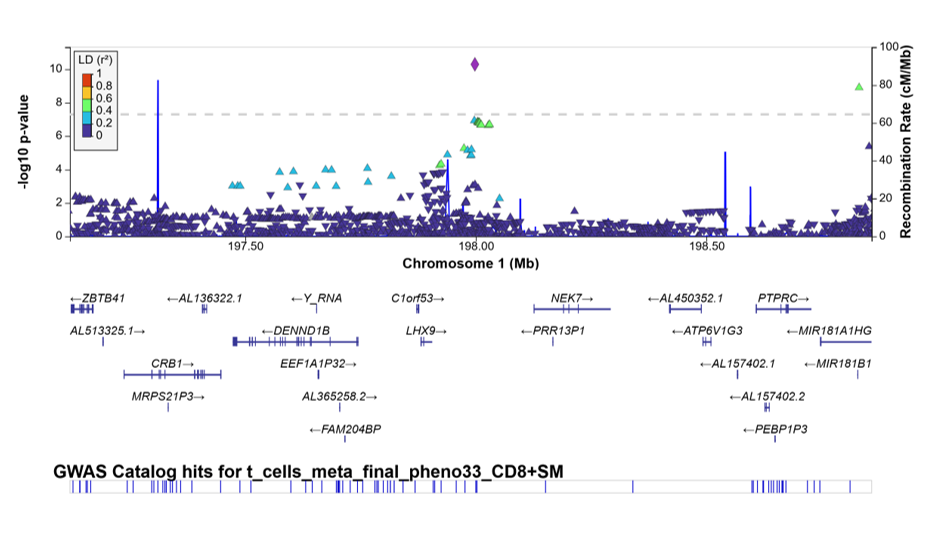

C. rs113116201 (*MIR181A1HG/PTPRC*; CD8+LD)

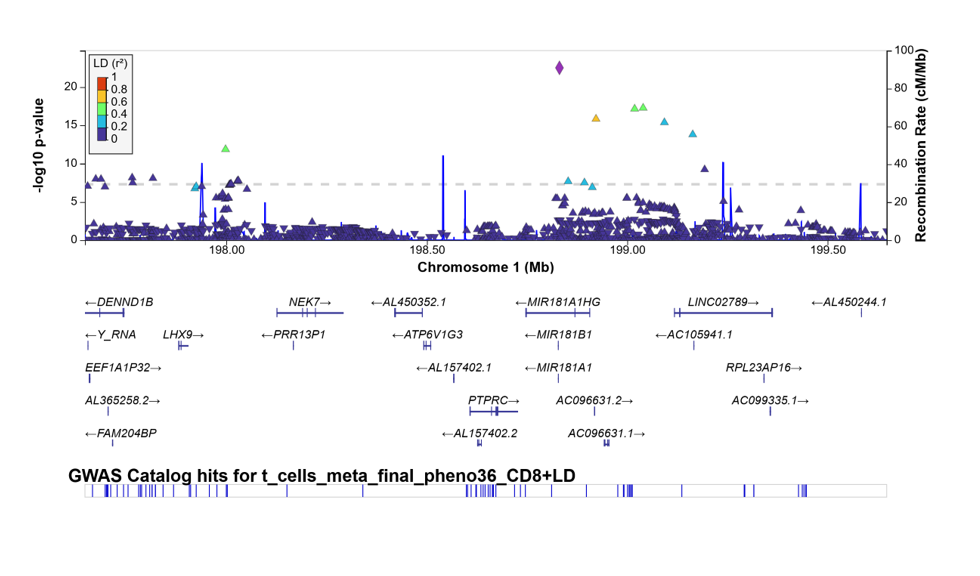

D. rs116517602 (*MIR181A1HG/PTPRC*; CD4+LEM)

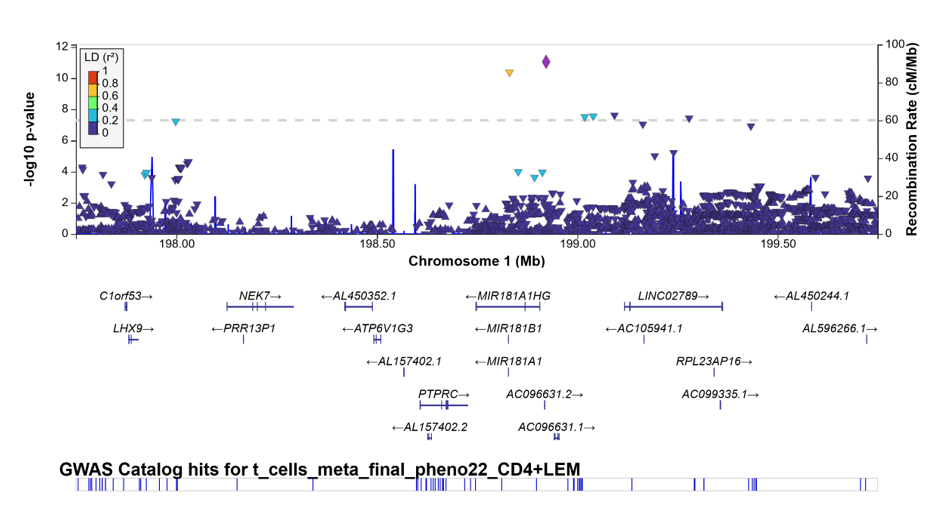

E. rs58905133 (*GALM*; CD8+CM_EM)

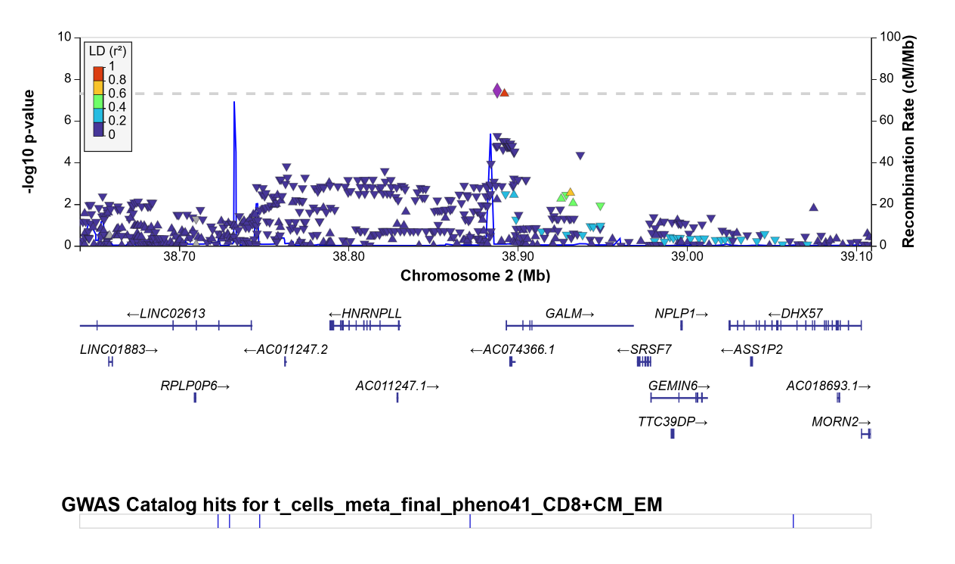

F. rs6809606 (*FHIT*; CD14+CD15-)

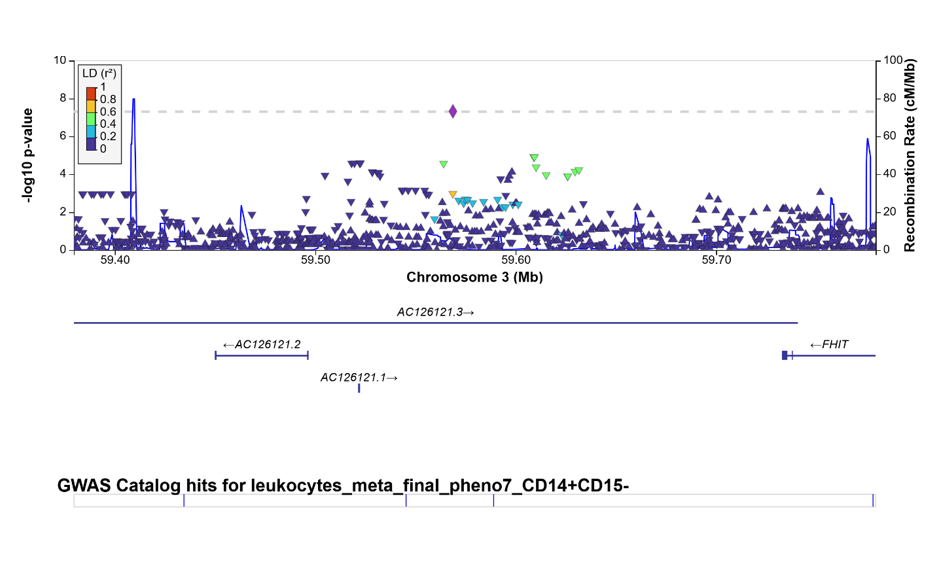

G. rs1878469 (*AC108142.1*; CD4+LEM)

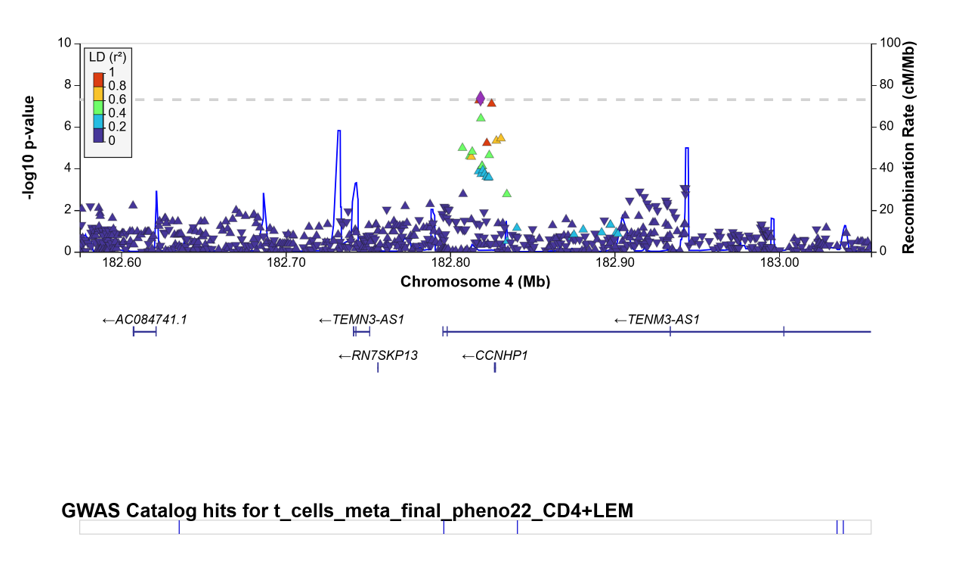

H. rs34356513 (*LMBRD2*; DC)

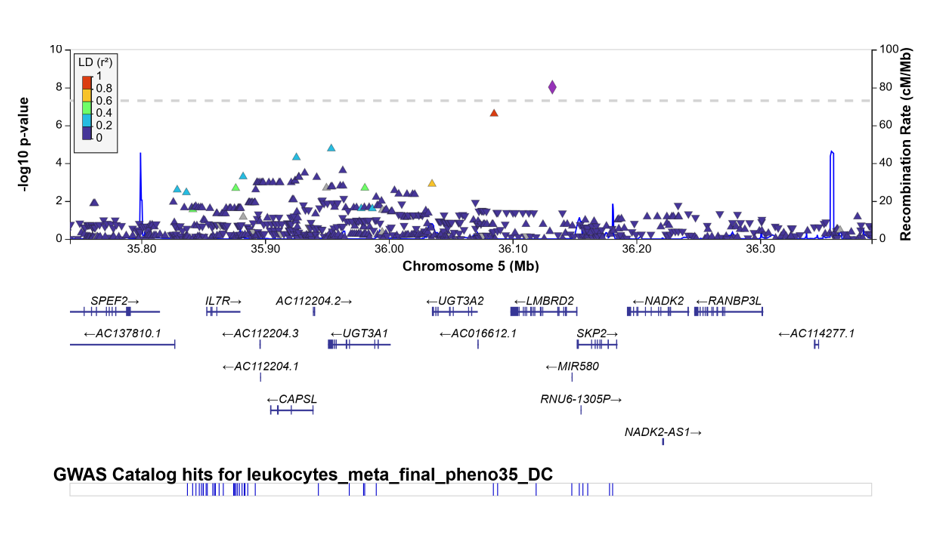

I. rs13159713 (*SLC1A3*; Tcell_NKG2C+)

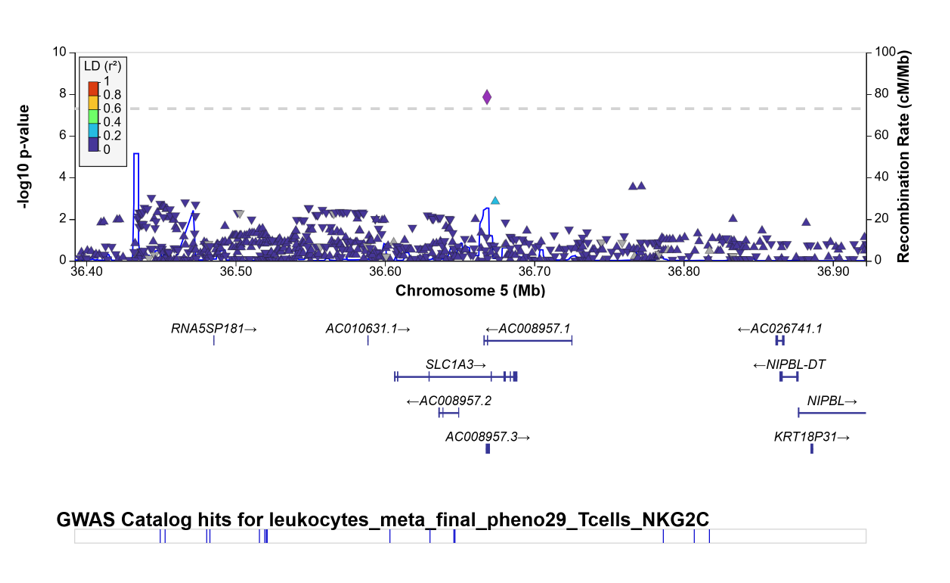

J. rs190088867 (*RP11-3N13.2*; CD4+LD)

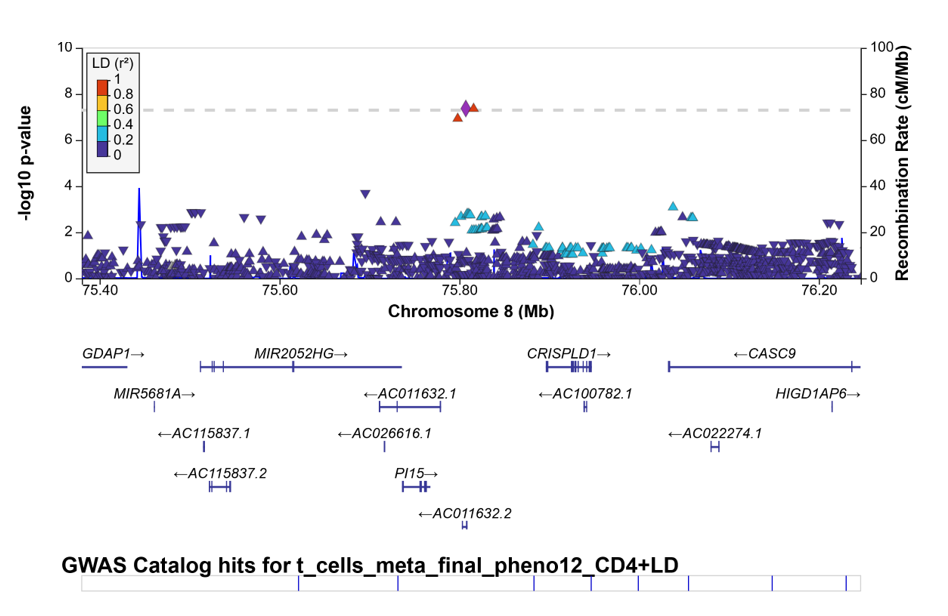

K. rs117904683 (*ZFHX4*; CD4+CM_EM)

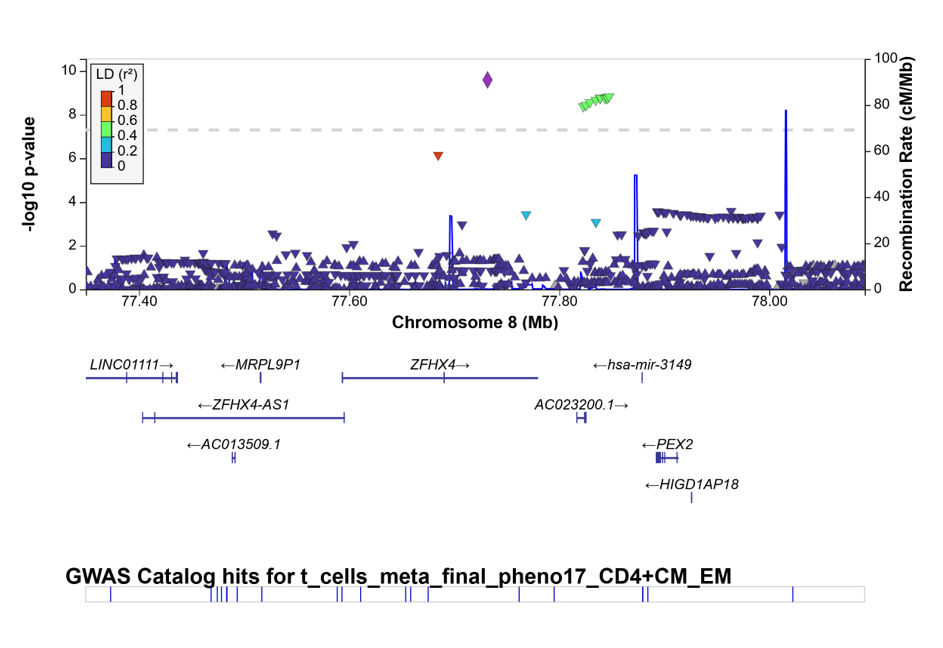

L. rs4739631 (*STMN2*; CD4+naive)

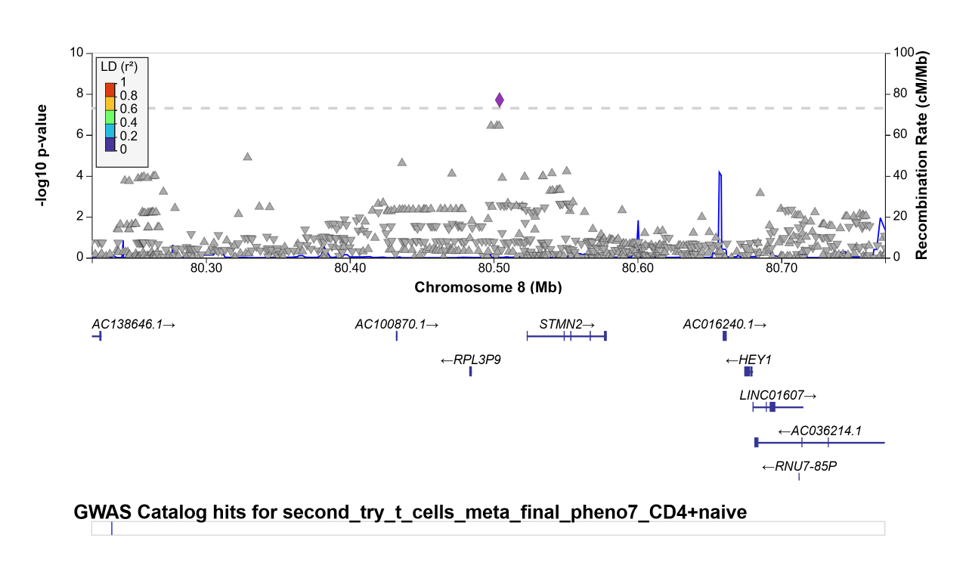

M. rs79059742 (*TBATA*; CD4/CD8 ratio)

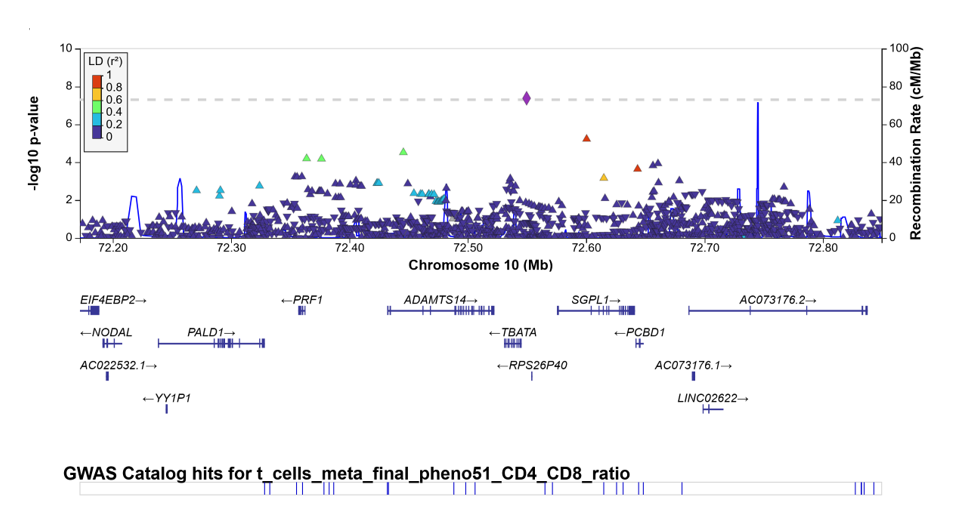

N. rs12257092 (*FAS*; CD4+SM)

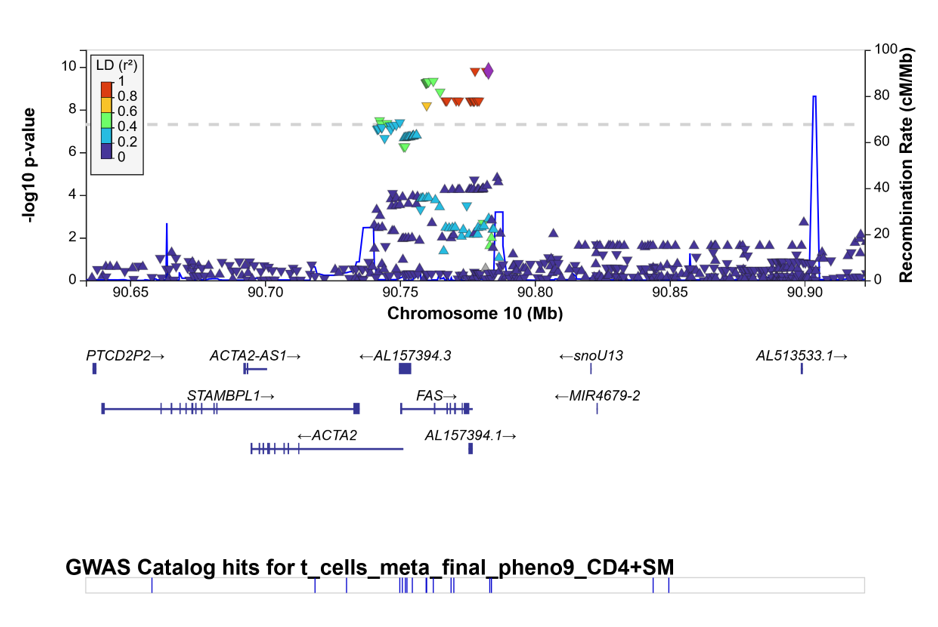

O. rs11822913 (*B3GAT1*; CD8+ME_CD57+)

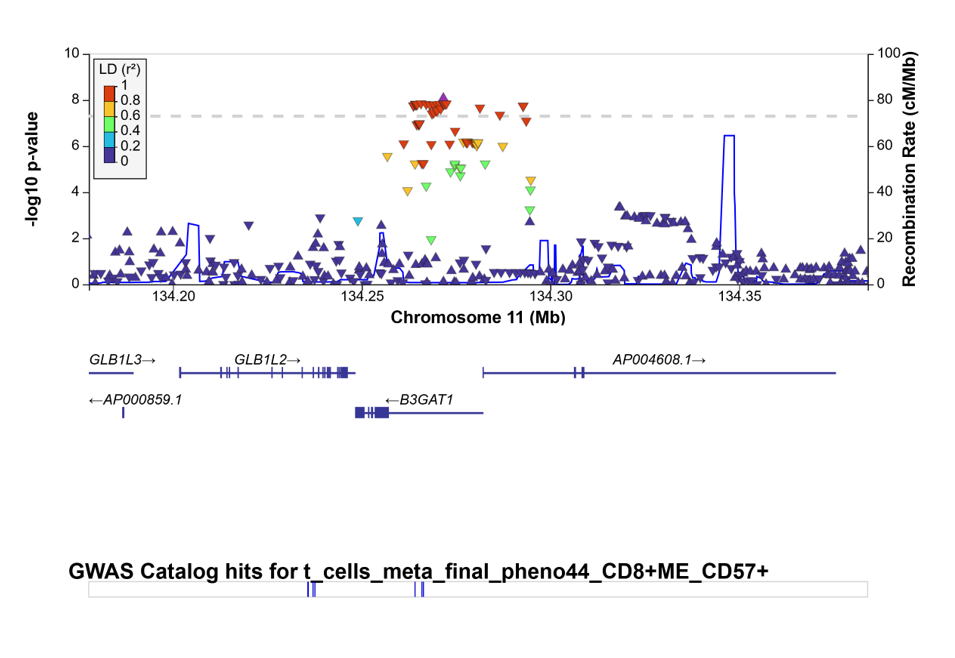

P. rs77753585 (*ANO2*; CD4+CD28+)

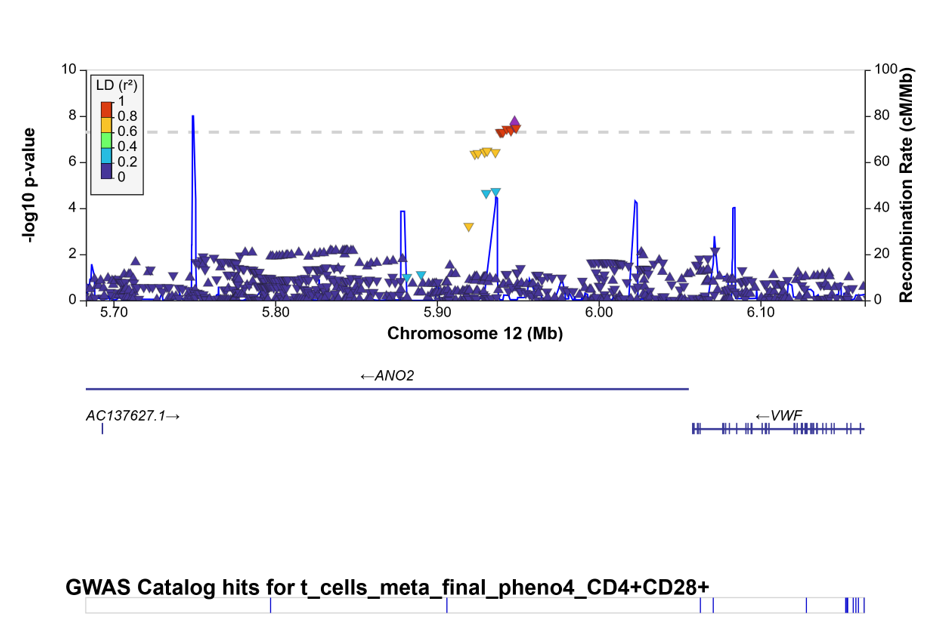

Q. rs12318583 (*KLRC3*; NK_immature_NKG2C+)

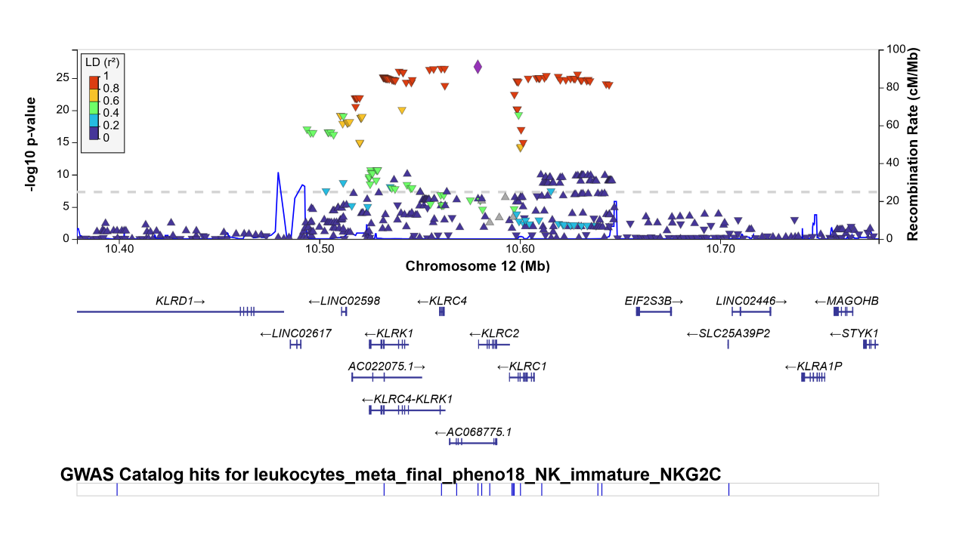

R. rs11053803 (*KLRC1*; NK_mature_NKG2A+)

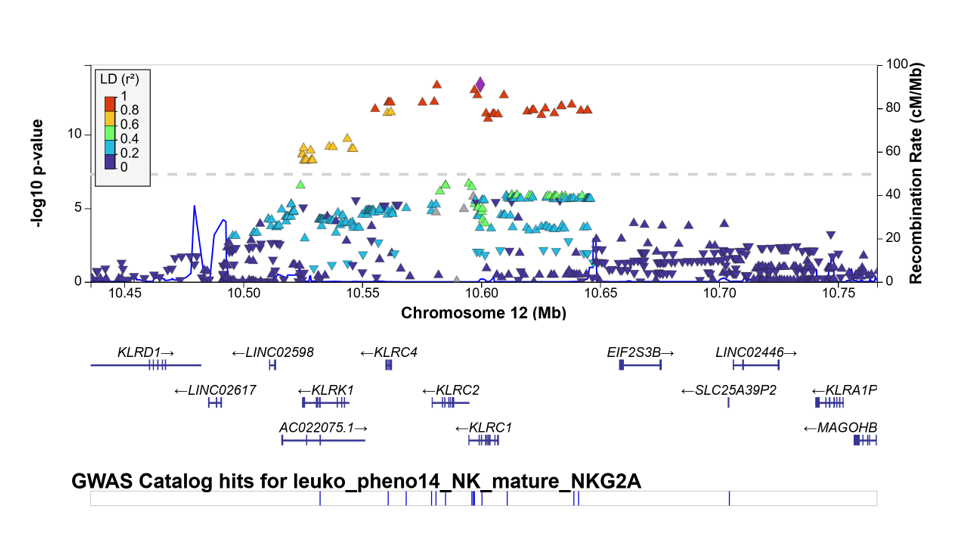

S. rs185320614 (*RBFOX1*; CD4+LD_CD27+)

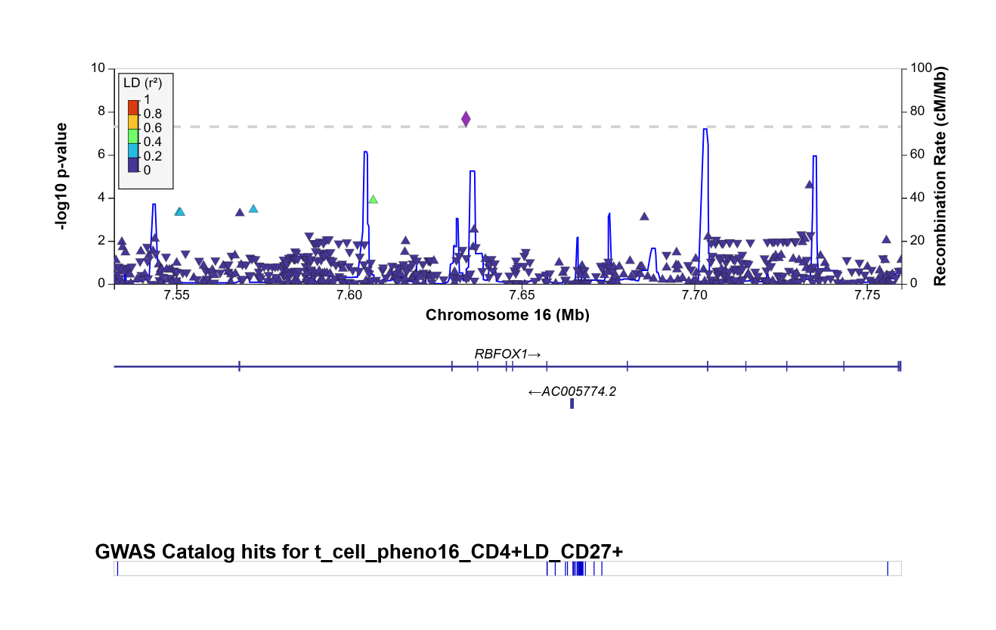

T. rs7225029 (*RNF213*; CD4+LD_CD27+CD28+)

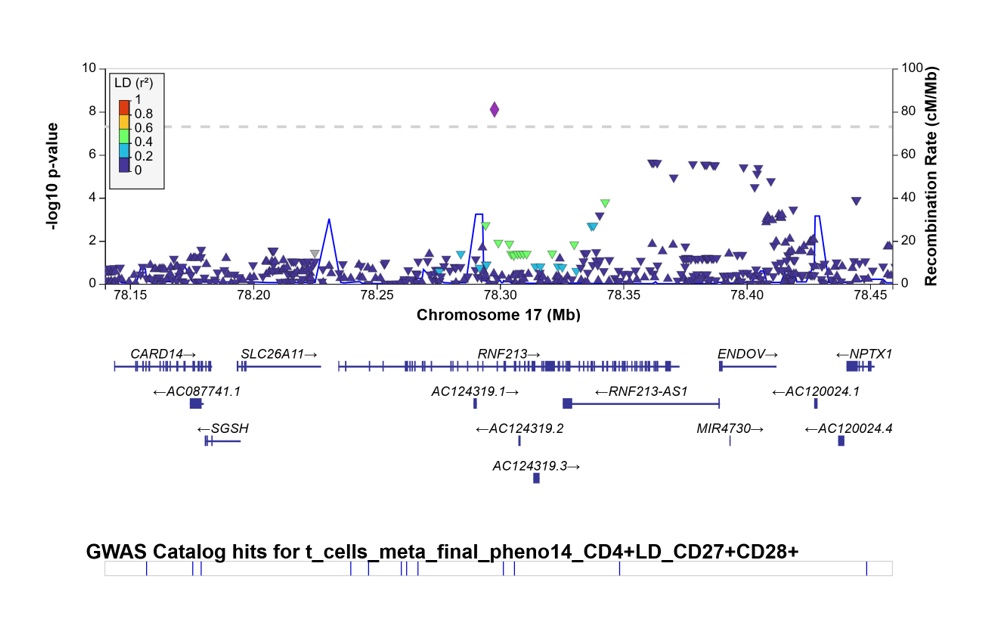

U. rs114460626 (*RMC1*; CD4+ME)

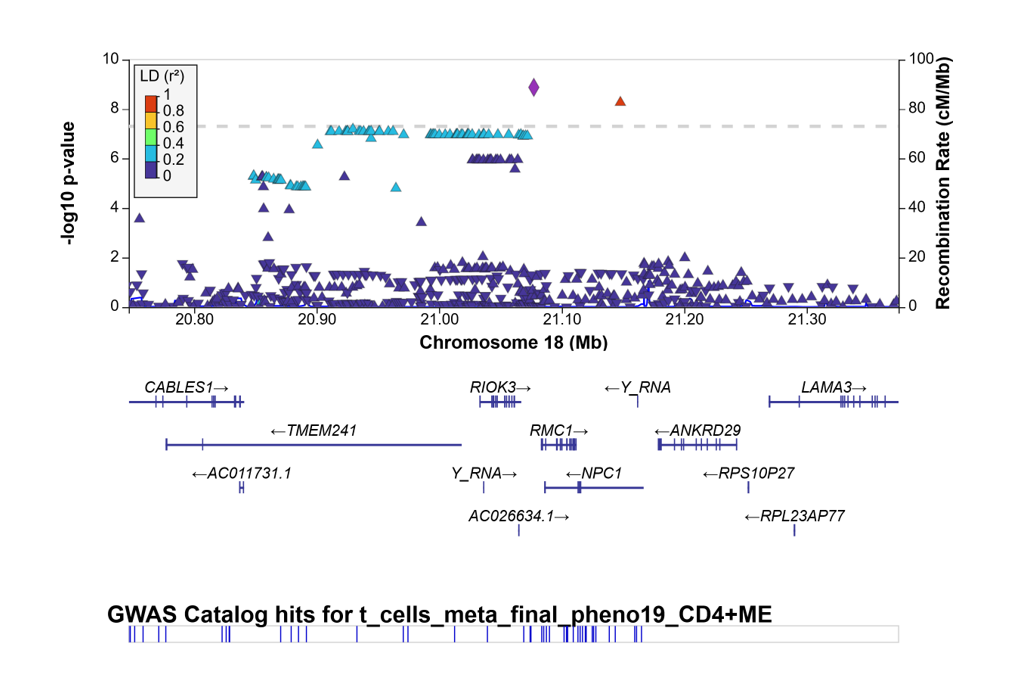

V. rs75060966 (*ST8SIA5*; MDSC4)

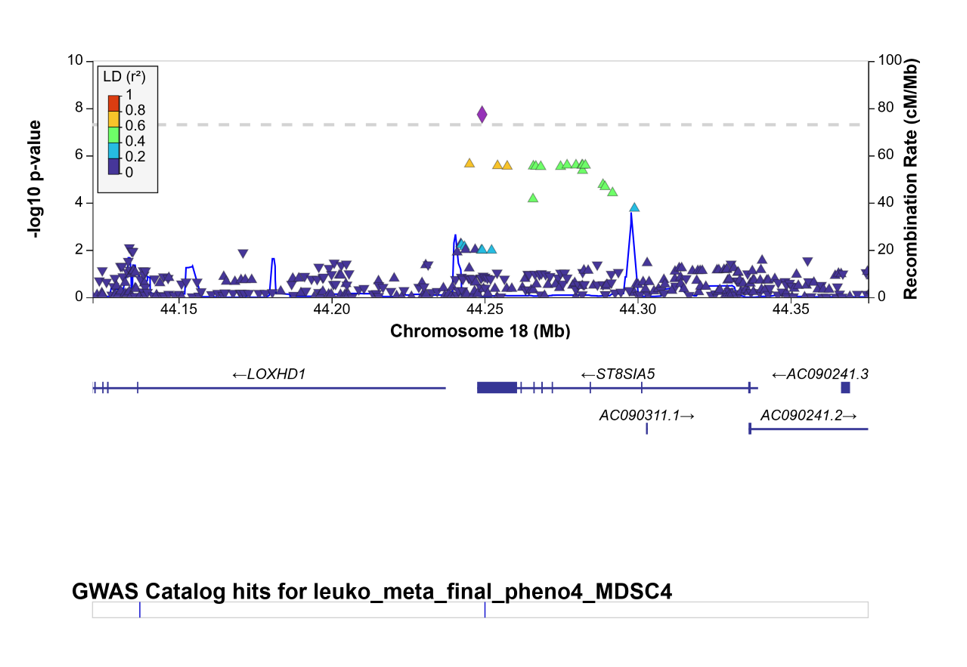

W. rs6077080 (*SLC52A3*; CD8+naive_CD57+)

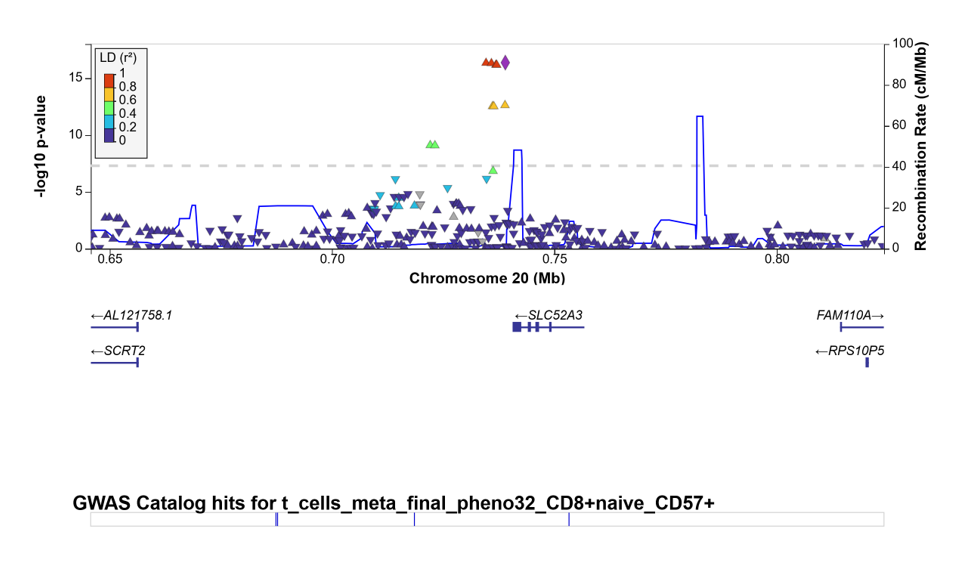

X. rs6116364 (*ADRA1D*; CD3-CD16-)

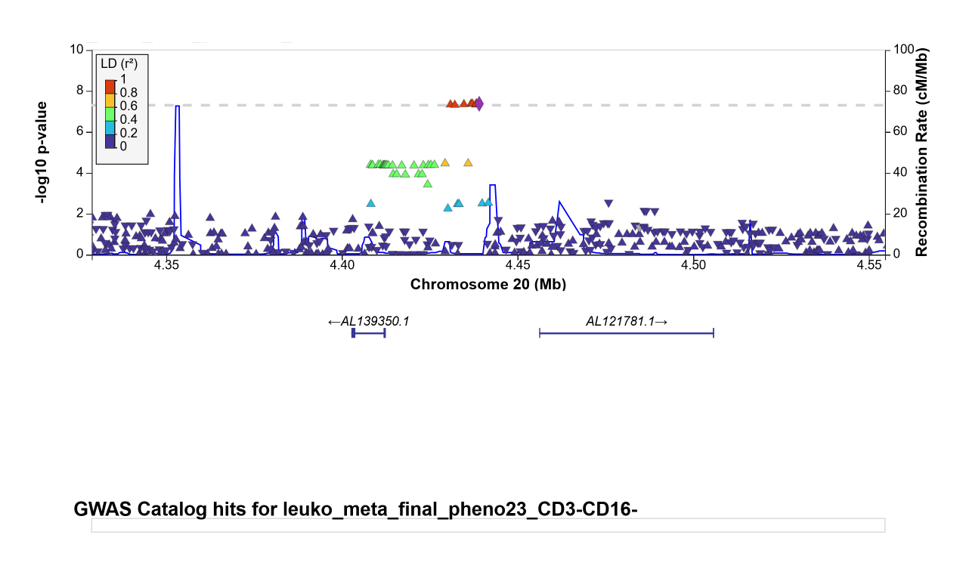

Legend. Locus zoom plots of genome-wide significant (α=5.00E-8) index SNPs of the immune cell GWAS in BASE-II. The sub headers provide the index SNP identifier (nearest gene; associated immune cell phenotype). The index SNP is visualized as a purple diamond. The color coding explained in the legend refers to the LD structure of all SNPs displayed with regard to the index SNP. These plots were produced with LocusZoom^32^ (reference population: “EUR”, all genes displayed [“all features”]).

**Supplementary References**

1.	Bertram, L. *et al.* Cohort Profile: The Berlin Aging Study II (BASE-II)†. *Int. J. Epidemiol.* **43**, 703–712 (2014).

2.	Lill, C. M. *et al.* Genetic Burden Analyses of Phenotypes Relevant to Aging in the Berlin Aging Study II (BASE-II). *Gerontology* **62**, 316–322 (2016).

3.	Sbierski-Kind, J. *et al.* T cell phenotypes associated with insulin resistance: results from the Berlin Aging Study II. *Immun. Ageing* **17**, (2020).

4.	Deecke, L. *et al.* No increase of CD8+ TEMRA cells in the blood of healthy adults at high genetic risk of Alzheimer’s disease. *Alzheimer’s Dement. J. Alzheimer’s Assoc. [in Press.* (2023).

5.	Charrad, M., Ghazzali, N., Boiteau, V. & Niknafs, A. NbClust : An R Package for Determining the Relevant Number of Clusters in a Data Set. *J. Stat. Softw.* **61**, (2014).

6.	Hong, S. *et al.* Genome-wide association study of Alzheimer’s disease CSF biomarkers in the EMIF-AD Multimodal Biomarker Discovery dataset. *Transl. Psychiatry* **10**, (2020).

7.	Chang, C. C. *et al.* Second-generation PLINK: rising to the challenge of larger and richer datasets. *Gigascience* **4**, (2015).

8.	Auton, A. *et al.* A global reference for human genetic variation. *Nature* **526**, (2015).

9.	Fisher, R. A. *Statistical methods for research workers*. (Oliver and Boyd, 1934).

10.	Watanabe, K., Taskesen, E., van Bochoven, A. & Posthuma, D. Functional mapping and annotation of genetic associations with FUMA. *Nat. Commun.* **8**, (2017).

11.	Devlin, B. & Roeder, K. Genomic Control for Association Studies. *Biometrics* **55**, 997–1004 (1999).

31.	Choi, S. W. & O’Reilly, P. F. PRSice-2: Polygenic Risk Score software for biobank-scale data. *Gigascience* **8**, (2019).

32.	Boughton, A. P. *et al.* LocusZoom.js: interactive and embeddable visualization of genetic association study results. *Bioinformatics* **37**, 3017–3018 (2021).
